# Impact and cost-effectiveness of the 6-month BPaLM regimen for rifampicin-resistant tuberculosis: a mathematical modeling analysis

**DOI:** 10.1101/2023.07.28.23293104

**Authors:** Lyndon P. James, Fayette Klaassen, Sedona Sweeney, Jennifer Furin, Molly F. Franke, Reza Yaesoubi, Dumitru Chesov, Nelly Ciobanu, Alexandru Codreanu, Valeriu Crudu, Ted Cohen, Nicolas A. Menzies

## Abstract

**Background:** Emerging evidence suggests that shortened, simplified treatment regimens for rifampicin-resistant tuberculosis (RR-TB) can achieve comparable end-of-treatment outcomes to longer regimens. We compared a 6-month regimen containing bedaquiline, pretomanid, linezolid, and moxifloxacin (BPaLM) to a standard of care strategy using a 9- or 18-month regimen depending on whether fluoroquinolone resistance (FQ-R) is detected on Drug Susceptibility Testing (DST).

**Methods and Findings:** The primary objective was to determine whether 6 months of BPaLM is a cost-effective treatment strategy for RR-TB. We used genomic and demographic data to parameterize a mathematical model estimating long-term health outcomes measured in quality-adjusted life years (QALYs) and lifetime costs in 2022 USD ($) for each treatment strategy for patients 15 years and older diagnosed with pulmonary RR-TB in Moldova, a country with a high burden of TB drug resistance. For each individual, we simulated the natural history of TB and associated treatment outcomes, as well as the process of acquiring resistance to each of 12 anti-TB drugs. Compared to the standard of care, 6 months of BPaLM was cost-effective. It was estimated to reduce lifetime costs by $3,366 (95% UI: [1465, 5742] p<0.001) per individual, with a non-significant change in QALYs (−0.06; 95% UI: [-0.49, 0.032] p=0.790). For those stopping moxifloxacin under the BPaLM regimen, continuing with BPaL plus clofazimine (BPaLC) provided more QALYs at lower cost than continuing with BPaL alone. 6 months of BPaLM had at least a 93% chance of being cost-effective, so long as BPaLC was continued in the event of stopping moxifloxacin. 6 months of BPaLM reduced the average time spent with TB resistant to amikacin, bedaquiline, clofazimine, cycloserine, moxifloxacin and pyrazinamide, while it increased the average time spent with TB resistant to delamanid and pretomanid. Sensitivity analyses showed 6 months of BPaLM to be cost-effective across a broad range of values for the relative effectiveness of BPaLM, and the proportion of the cohort with FQ-R. Compared to the standard of care, 6 months of BPaLM would be expected to save Moldova’s national TB program budget $7.1 million (95% UI: [1.3 million, 15.4 million] p=0.002) over the five year period from implementation. This analysis did not account for all possible interactions between specific drugs as they apply to treatment effectiveness, to resistance acquisition, or to the consequences of specific types of severe adverse events, nor did it model how the intervention may affect the transmission dynamics of RR-TB.

**Conclusions:** Compared to the standard of care, the implementation of the 6-month BPaLM regimen could improve the cost-effectiveness of care for individuals diagnosed with RR-TB, particularly in settings where current long-course regimens are challenging to implement and afford. Further research may be warranted to explore the suitability of shorter RR-TB regimens in specific national settings.

**AUTHOR SUMMARY:** **Why was this study done?**

- Drug resistance poses a major barrier to the effective treatment of tuberculosis, especially in Moldova and other post-Soviet states which have the highest levels of resistance in the world.
- Individuals with tuberculosis resistant to the key drug rifampicin face a worse prognosis, a longer and more expensive course of treatment, and more side effects than individuals with rifampicin-susceptible tuberculosis.
- Until recently, the standard of care for rifampicin-resistant tuberculosis involved many drugs in combination, often given for 18 months or longer.
- The newer, 6-month “BPaLM” regimen is comprised of four drugs (bedaquiline, pretomanid, linezolid, moxifloxacin) to which resistance levels are currently low, and while it was shown to be just as effective as the standard of care for 72-week health outcomes, its effect on lifetime health outcomes, costs, and the acquisition of drug resistance was less clear.

**What did the researchers do and find?**

- Using a mathematical model, we projected the lifetime health benefits and costs of the 6-month BPaLM regimen as compared to the standard of care treatments for rifampicin-resistant tuberculosis, and found that 6 months of BPaLM provided similar health benefits to longer regimens, at lower cost.
- Compared to the standard of care, we also found that the 6-month BPaLM regimen shortened the average duration of tuberculosis that was resistant to the drugs amikacin, bedaquiline, clofazimine, cycloserine, moxifloxacin, and pyrazinamide, while it increased the average duration of tuberculosis resistant to delamanid and pretomanid.
- For individuals receiving BPaLM who had to stop taking the drug moxifloxacin, we found that it would be beneficial on both health and cost grounds to replace it with clofazimine, thereby topping the regimen back up to four drugs.

**What do these findings mean?**

- Using conventional benchmarks for value-for-money, 6 months of BPaLM is a cost-effective approach for the treatment of rifampicin-resistant tuberculosis in Moldova, and potentially other post-Soviet countries.
- Though the effect of the 6-month BPaLM regimen on the spread of drug resistance in the population is uncertain and not addressed directly by this study, this combination of newer drugs appears to achieve cure more quickly, which reduces the amount of time an individual is potentially infectious and so may be beneficial in fighting resistance to several drugs, even while it may increase the spread of resistance to others.
- Further studies may be warranted to explore how well these findings translate to different global regions where health system capabilities, costs, and existing resistance patterns may differ.

## INTRODUCTION

Treatment for rifampicin-resistant tuberculosis (RR-TB) is complex, involving combinations of several drugs—many of which have substantial potential for toxicity—over a prolonged course of therapy. The 2022 WHO Guidelines for the treatment of drug-resistant tuberculosis recommend a shorter, 6-month regimen composed of bedaquiline, pretomanid, linezolid, and moxifloxacin (BPaLM) to treat rifampicin-resistant tuberculosis (RR-TB) [1]. These guidelines updated earlier 2020 WHO Guidelines that recommended several treatment regimens, each comprising 4-7 drugs for 9-18 months or longer [2].

The evidence base for shorter regimens for RR-TB has been broadly positive, including results from observational studies [3,4], single-arm clinical trials [5,6], mathematical modeling analyses [7], and the recent multicenter open-label randomized controlled trial TB-PRACTECAL [8]. Although trial recruitment was stopped early on the recommendation of a planned, interim review by the study monitoring committee, the analysis suggested that 6 months of BPaLM was non-inferior to the standard of care with respect to treatment outcome (a composite of death, treatment failure, treatment discontinuation, loss to follow-up, or recurrence) and was beneficial with respect to safety [8]. The adoption of shorter, simplified regimens may be further bolstered by the forthcoming publication of the results of the endTB trial [9–13], but the absence of larger, confirmatory trials led to a conditional recommendation by the WHO in 2022. The pursuit of effective shorter treatment regimens is also driven by the desire to alleviate the considerable psychological and emotional toll of prolonged treatment for RR-TB. On top of drug side effects [14], many patients undergoing treatment for RR-TB experience stigma, depression, loss of self-esteem, and economic hardship from an inability to work [15]. Patients may lack access to sufficient psychological and financial supports [16–18], and this may be particularly hard for individuals with housing or employment instability, or substance use disorder [19].

The 2020 WHO Guidelines represent the existing standard of care in many settings. In addition to higher prices and supply constraints for newer drugs [20],[21], it is expected that the rollout of the BPaLM regimen as part of the newer 2022 Guidelines may be delayed by concerns about comparative effectiveness and cost-effectiveness [20–24]. Implementation may also be met with concern over the emergence of drug resistance, particularly in settings with limited capacity to detect resistance to newer agents such as bedaquiline, pretomanid, and linezolid [25]; such capacity constraints are multifactorial, from the expense of investing in new technologies and associated laboratory workforce development, to supply chain interruptions and divergent political priorities [26,27]. The decision to implement the new 6-month BPaLM regimen will depend on setting-specific tradeoffs between regimen effectiveness, cost, the complexity of treatment decisions, and existing levels of resistance to anti-TB drugs in the population. Decision analysis provides a framework to analyze these tradeoffs, and a recent cost-effectiveness study using evidence from TB-PRACTECAL found that 6-months BPaLM may reduce cost and improve health relative to the standard of care in several countries [28]. Our analysis builds on this work by focusing on longer term outcomes that are difficult to measure in a trial setting and by examining a wider range of testing and treatment approaches, including whether patients who must stop moxifloxacin—due to side effects or acquired resistance—should continue on BPaL alone, or BPaL plus clofazimine (BPaLC) [25,28].

In this study, we investigated the health impact and cost-effectiveness a 6-month BPaLM regimen for the treatment of adults with pulmonary RR-TB, as compared to the standard of care. We considered a range of treatment strategies incorporating these two approaches, varying the timing and frequency of drug susceptibility testing (DST) as well as how regimens would be modified for individuals developing fluoroquinolone resistance. To estimate outcomes, we used a Markov microsimulation model parameterized with detailed genomic sequencing data describing specific patterns of initial drug resistance, and calculated the effect of each treatment strategy on length and quality of life as well as costs, accounting for regimen effectiveness, risks of severe adverse events (SAEs) due to drug toxicity, and acquisition of resistance.

We conducted the analysis for the setting of Moldova, an upper-middle income post-Soviet country where the incidence rate of RR-TB is among the highest in the world, and where an estimated 33% of individuals newly diagnosed with TB have RR-TB, ten times higher than the same proportion globally [29,30]. The reasons for this picture are not fully understood, but it is thought that economic shocks following the breakup of the Soviet Union contributed to this picture in the region, along with early treatment discontinuation [31] and mass incarceration [32]. In recent years in Moldova, a multidisciplinary committee reviews the treatment course of every patient receiving treatment for RR-TB, and WHO treatment guidelines are closely adhered to [VC, DC]. Moldova also has developed a robust TB laboratory infrastructure, which provided a platform for recent genomic sequencing of culture-positive isolates [33]. By harnessing this genomic resistance data, we hope to improve the cost-effectiveness of treatment in a country with a very high burden of RR-TB, as well as other countries in the region. We also explored the generalizability of our findings to settings with a different prevalence of initial fluoroquinolone resistance among RR-TB.

## METHODS

### Strategies

We compared eight treatment strategies, each reflecting a different approach to drug regimen choice and timing of DST (Table 1). Two strategies adopted drug regimens aligned with the standard of care as defined by the 2020 WHO Guidelines [2], with all individuals started on a WHO longer regimen while awaiting the results of second-line DST by mycobacterial growth indicator tube (MGIT) to fluoroquinolones and injectables. Fluoroquinolone resistance (FQ-R) identified via MGIT was assumed to result in the continuation of an 18-month WHO longer regimen, with refinements as necessary based on DST. If fluoroquinolone susceptibility (FQ-S) was detected, treatment was switched to a 9-month regimen (S1 Fig). Under one standard of care strategy (strategy (7)), we modelled the minimum guideline-recommended frequency of second-line DST–every 4 months, and in another (strategy (8)) we increased this to a monthly frequency. While the 2020 WHO Guidelines did not prescribe exactly one combination of drugs for each scenario, we adopted a single combination of drugs for each situation for tractability, based on our best interpretation of the guideline’s hierarchy of group A, B, and C drugs (S1 Fig).

**Table 1.** Key features of the modeled RR-TB treatment strategies.

| Strategy No. | Guidelines informing the strategy | Drug regimen | Regimen duration | For BPaLM-based strategies only, prescribed regimen for those who discontinue Moxifloxacin | Replacement drugs, in order, for all other discontinuations | DST for second-line drugs (MGIT) at treatment initiation | Routine frequency of subsequent DST for second-line drugs | Indications for drug discontinuation | Length of regimen extension, if necessary* |
| --- | --- | --- | --- | --- | --- | --- | --- | --- | --- |
| 1 | 2022 WHO Guidelines | BPaLM | 6 months | BPaLC | Clofazimine, Cycloserine | Yes | 4 months | Immediately following:<br>• Resistance identified on DST<br>• Grade 4-5 Severe Adverse Event | 6 months |
| 2 | 2022 WHO Guidelines | BPaLM | 6 months | BPaLC |  | Yes | 1 month |  |  |
| 3 | 2022 WHO Guidelines | BPaLM | 6 months | BPaL |  | Yes | 4 months |  |  |
| 4 | 2022 WHO Guidelines | BPaLM | 6 months | BPaL |  | Yes | 1 month |  |  |
| 5 | 2022 WHO Guidelines | BPaLM | 6 months | BPaLC |  | No | 4 months |  |  |
| 6 | 2022 WHO Guidelines | BPaLM | 6 months | BPaL |  | No | 4 months |  |  |
| 7 | 2020 WHO Guidelines (standard of care) | Start treatment with WHO Longer regimen (bedaquiline, clofazimine, linezolid, moxifloxacin), await second-line DST.<br>**<u>If FQ-R</u>**, continue on WHO Longer regimen, i.e.: |  |  |  | Yes | 4 months |  |  |
|  |  | Bedaquiline, Clofazimine, Linezolid, Cycloserine | 18 months | n/a | Ethambutol, Delamanid, Pyrazinamide, Amikacin, Ethionamide |  |  |  |  |
|  |  | **<u>If FQ-S</u>**, switch to 2020 WHO shorter, all-oral bedaquiline containing regimen: |  |  |  |  |  |  |  |
|  |  | Bedaquiline, Clofazimine, Ethambutol, Ethionamide, Isoniazid, Moxifloxacin, Pyrazinamide | 9 months | n/a | Delamanid, Cycloserine |  |  |  |  |
| 8 | 2020 WHO Guidelines (standard of care) | As for Strategy No. 7 |  |  |  | Yes | 1 month |  |  |

BPaL – bedaquiline, pretomanid, linezolid; BPaLC – bedaquiline, pretomanid, linezolid, clofazimine; BPaLM – bedaquiline, pretomanid, linezolid, moxifloxacin; DST – drug susceptibility test; FQ-S – fluoroquinolone susceptible; FQ-R – fluoroquinolone resistant; MGIT – mycobacterial growth indicator tube; WHO – World Health Organization
\*Regimen extensions were implemented for those who had not yet successfully completed treatment. While the 2020 WHO Guidelines recommended the BPaL regimen in specific situations, none of the modeled cohort met the inclusion criteria to receive BPaL under those strategies.

The remaining six strategies were modeled on the 2022 WHO Guidelines [34] with 6-month BPaLM-based regimens. In three of these strategies, individuals having to stop Moxifloxacin (because of a SAE or because resistance was detected on DST) were continued on BPaL alone, as recommended by the 2022 Guidelines. In the remaining three, they continued on BPaLC. The remaining differences between these six strategies depended on the prescribed schedule of DST to second-line drugs; in two of these strategies, we explored the effects of omitting routine second-line DST at treatment initiation (Table 1).

### Population and data

We modeled a cohort of individuals aged 15 years and older diagnosed with RR-TB in Moldova. For each individual, their age and the resistance profile of the strain of *M. tuberculosis* causing infection were informed by publicly available genomic sequencing data from Moldova [35]. These data comprised single-strain *M. tuberculosis* samples collected in 2018–2019; a full description has been provided by Yang and colleagues [30]. We assumed that a mutation associated with resistance conferred full resistance to that drug. Conversely, *M. tuberculosis* strains lacking relevant resistance mutations were assumed to be fully susceptible to the respective drugs. We excluded data for rifampicin susceptible strains (S2 Fig) leaving 674 distinct samples from which we simulated the modeled population. The proportion of isolates with resistance to each drug is shown in S3 Fig. This analysis used publicly available data only, and did not require ethical approval.

### Model

We used a Markov microsimulation model to simulate lifetime outcomes for a cohort of 10,000 individuals. Individuals in the model were simulated by random draws from the genomic sequencing dataset, with replacement. They were each assigned a drug regimen based on the modeled strategy (Table 1). Individuals then were assumed to transition between four health states: (1) Receiving TB treatment, (2) TB disease – not receiving treatment, (3) Cured post-treatment, and (4) Dead (S4 Fig). Within each Markov state, individual events were tracked including true cure as a result of treatment or self-cure, the occurrence of SAEs, second-line DST, changes to the drug regimen, loss to follow-up, relapse, death, and the evolution of drug resistance for that individual’s strain of *M. tuberculosis*. Extensions to the treatment regimen were implemented for those not observed to have successfully completed treatment.

While the range of SAEs resulting from TB treatment are of many varying durations and degrees of impact on quality of life, we accounted for these events in a simplified way by modeling the risk of a grade 4-5 SAE during the first three months of exposure to each drug, with each grade 4-5 SAE conferring a small but lifelong deduction in quality of life (Table 2, S1 Table). Grade 4-5 SAEs and diagnosed resistance constituted lifetime contraindications to the relevant drug, and replacements were made according to the modeled strategy (Table 1).

**Table 2.**
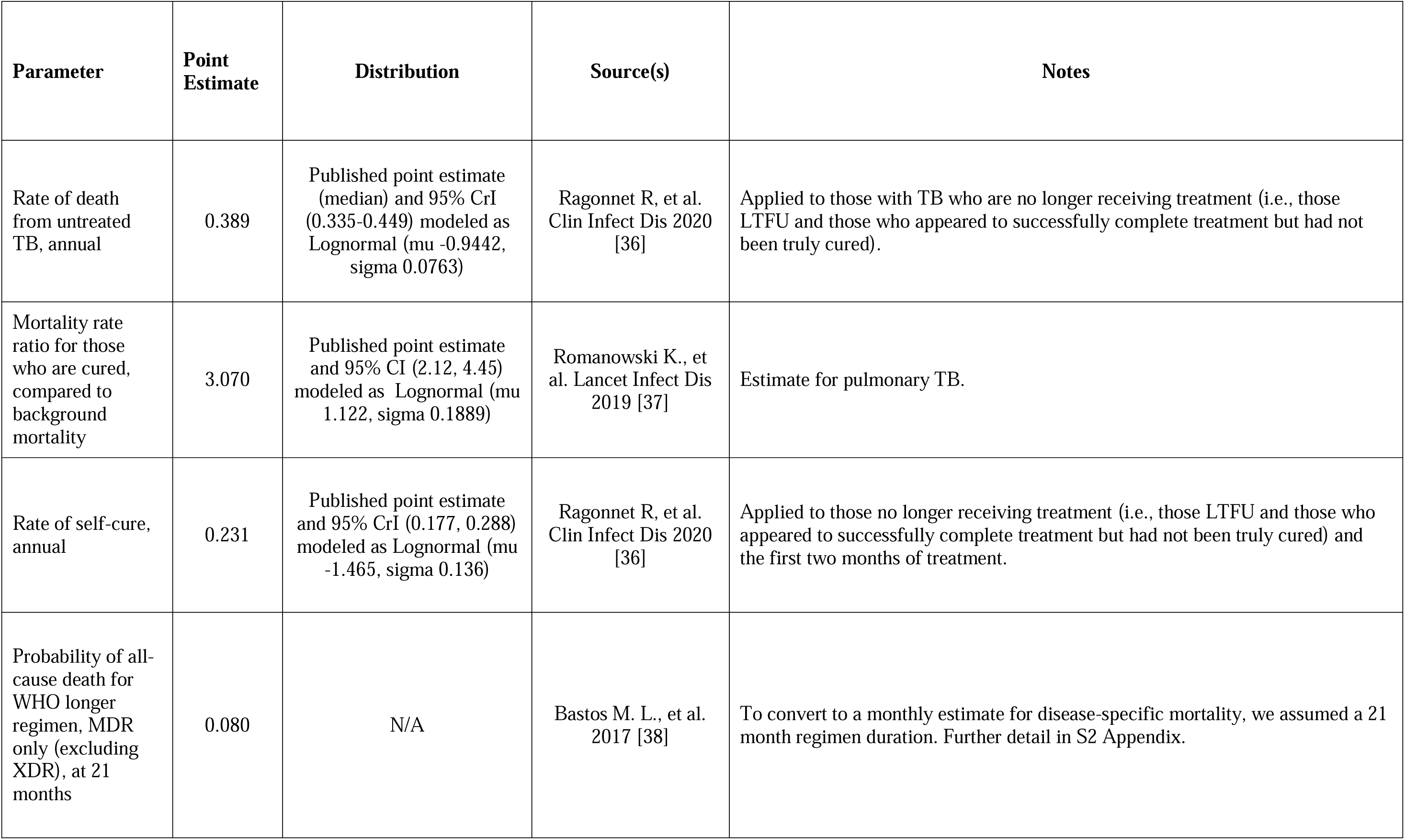
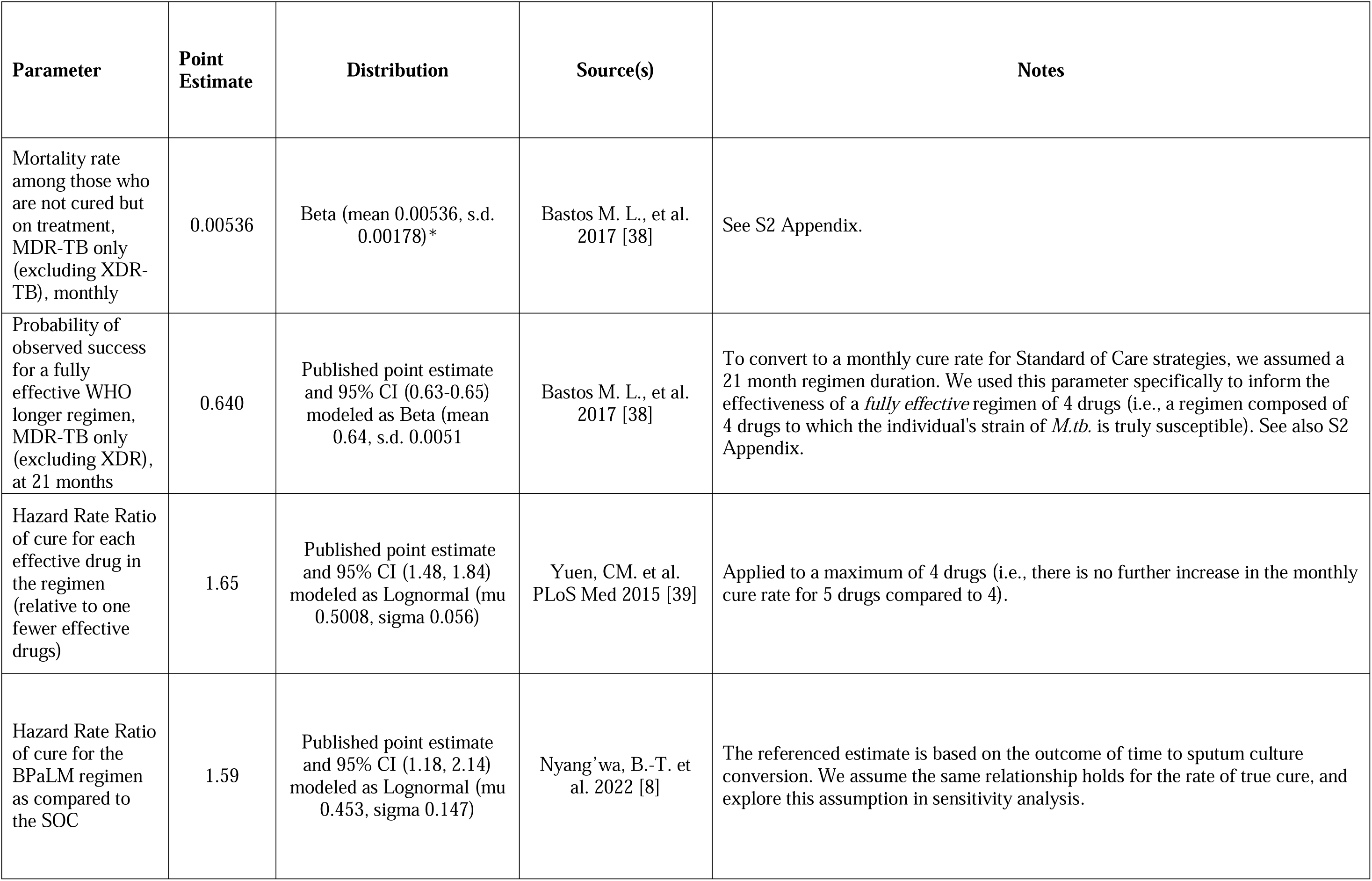
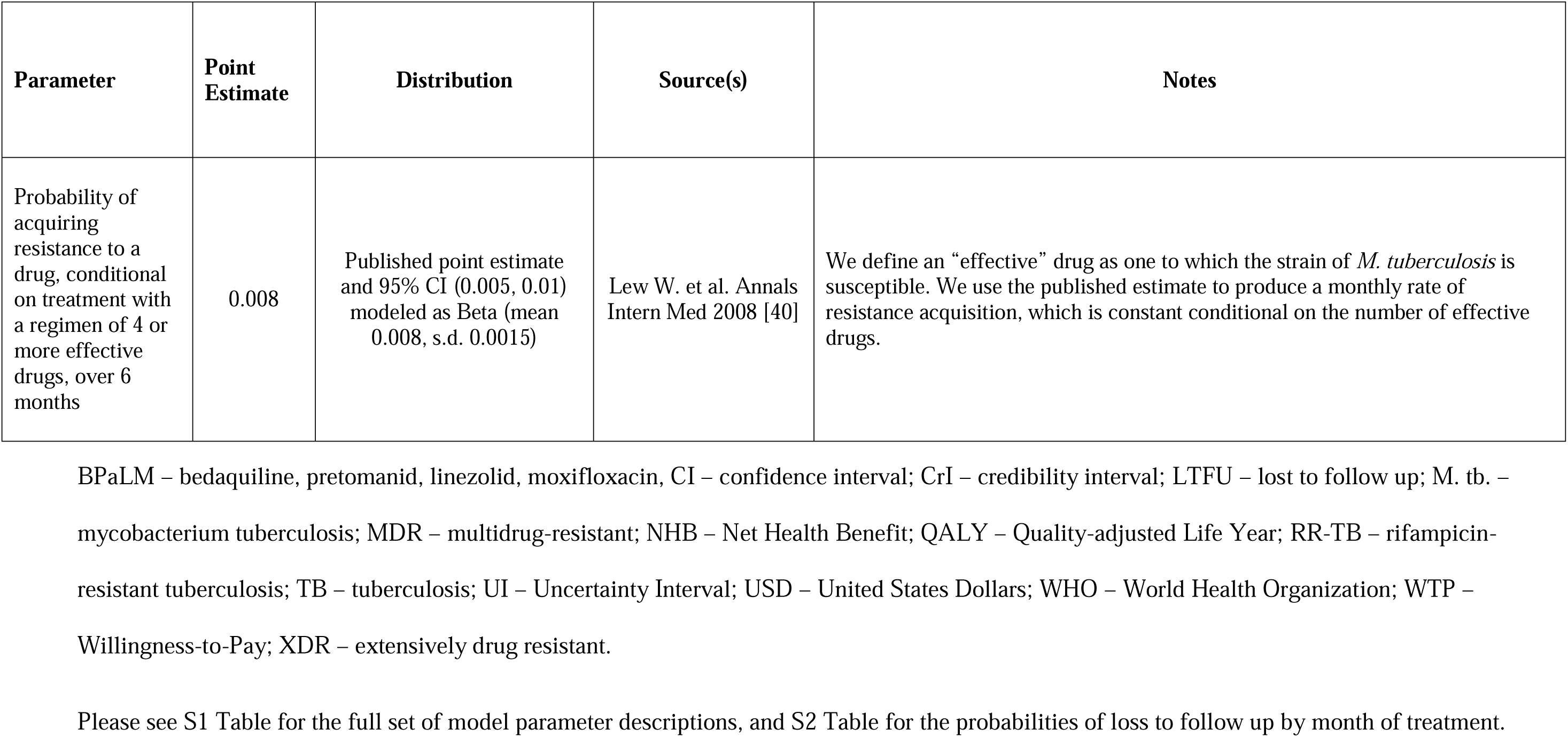
Key model parameters.

Each month, we tracked the drug regimen and the true resistance profile of each individual’s strain of *M. tuberculosis*. The number of effective drugs in a regimen was defined as the sum of all drugs being received, minus those drugs to which the strain of *M. tuberculosis* was resistant. The effect estimate for cure in BPaLM-based strategies as compared to the standard of care was modeled as the trial estimate for sputum culture conversion from TB-PRACTECAL, conditional on the number of effective drugs in the regimen, up to a maximum of four (i.e., four effective drugs confer a higher monthly cure rate than three, but five or more effective drugs do not confer a higher monthly cure rate than four) [8]. We varied this parameter in sensitivity analysis. S3 Fig displays the modeled rate of acquisition of new resistance to each drug, which was also conditioned on the number of effective drugs, to a maximum of four. S1 Table details the derivation and values for these and all other model parameters. DST was performed at a frequency informed by the strategy (Table 1), with sensitivity and specificity incorporated for each (S1 Table). Additional detail on model structure is provided in the S1 Appendix.

### Outcomes

The primary health outcome was measured in quality-adjusted life years (QALYs), a conventional approach in cost-effectiveness analysis [41,42]. QALYs measure the total length of time lived, weighted by time-varying, health-related quality of life, where 1 QALY is valued equivalently to one year in a state of perfect health [42,43]. This approach integrates the effects of the treatment strategies on both length and quality of life. QALYs rely on several assumptions, including risk-neutrality over length of life, and that maximizing total population QALYs—summed across all individuals—is desirable [44]. For each modeled individual in each month, we assigned health-related quality of life weights on a scale from 0 (dead) to 1 (perfect health), and multiplied the weight by 1/12 to account for the one-month cycle length (i.e., a dead individual accrued 0 QALYs for that month, and an individual in perfect health accrued 1/12 QALYs). These estimates were lowest on average at presentation and increased during the course of treatment; grade 4-5 SAEs also conferred a decrement in the utility weight (S1 Table). The total QALYs were calculating by summing all the month-specific QALYs accrued over each modeled individual’s lifetime.

We measured the impact on drug resistance by summing for each individual, and for each of 12 anti-TB drugs, the number of months they experienced TB disease with resistance to that drug. We then calculated three summary measures for the impact on drug resistance. In the first, we calculated the mean duration with resistance to each drug for the entire cohort by aggregating the time with resistance across the whole cohort for each drug, then dividing by the size of the starting cohort. Second, we calculated the mean duration of *untreated* TB disease with resistance to each drug by summing the time with resistance only among those individuals in Markov state (2)—TB disease no longer receiving treatment—and again averaging across the starting cohort. These measures were designed to reflect the relevance of the policies for the transmission of drug resistance. We calculated both because—for individuals no longer receiving treatment—there could be a higher risk that *M. tuberculosis* will transmit to another host, compared to the cohort as a whole. Third, we calculated the lifetime cumulative incidence of acquiring resistance to each drug, per individual in the cohort.

As a set of secondary health outcomes, we calculated the number of grade 4-5 SAEs experienced per patient to each of the drugs, and total life years (LYs, i.e. not weighted by health-related quality of life). To permit the validation of our model results, we also tracked two types of shorter-term outcomes at 6 months, 12 months, and 17 months (i.e., 72 weeks, the trial endpoint in TB-PRACTECAL): a) the proportion of individuals experiencing the end-of-treatment outcomes of Success, Failed by Treatment, Lost to Follow-up (LTFU), and Dead, as would typically be reported programmatically to the WHO; and b) a composite unfavorable outcome, including Death, LTFU, failed by treatment, and grade 4-5 SAEs, based on the primary outcome in TB-PRACTECAL [8].

We measured the total costs under each strategy from a societal perspective in 2022 United States dollars ($) as the sum of direct medical, direct non-medical, and indirect costs accruing in each period. Direct medical costs (i.e., those arising directly from the consumption of healthcare goods and services) were calculated by adding the costs of the drugs received, laboratory culture and DST to second-line drugs, a baseline healthcare resource utilization in the form of inpatient and outpatient services, and the cost of LTFU tracing. Direct non-medical and indirect costs were informed by published estimates for Moldova [45]. Each grade 4-5 SAE was accompanied by a utilization cost for inpatient and outpatient services. Direct non-medical costs (e.g., transportation) and indirect costs (e.g., productivity losses) accrued for every additional month on treatment. The indirect costs also accrued for those LTFU prior to cure. Productivity losses secondary to early mortality were not included in total costs, and were calculated separately.

Undiscounted values were calculated for all outcomes. For QALYs and total costs only, discounted values were also calculated using an annual discount rate of 3%.

#### Cost-effectiveness analysis

First, we ruled out dominated strategies (i.e., those strategies that were both more expensive and provided fewer QALYs on average than a linear combination of other strategies. We then calculated the relevant incremental cost-effectiveness ratios (ICER; a measure of the additional cost required to produce one additional QALY, as compared to the next cheapest, non-dominated strategy). We identified the cost-effective strategy as that with the greatest health gains, subject to the constraint that— in order to provide value for money—the ICER must be below the willingness-to-pay (WTP) threshold [41,46]. Lower ($4700 per QALY) and higher ($7021 per QALY) benchmarks for these thresholds in Moldova were based on published estimates using an opportunity cost approach [47], updated to 2022 USD (S1 Table). As ICERs may be challenging to interpret in some cases [48], we also calculated the Net Health Benefit (NHB) of each strategy (see S1 Appendix), with the cost-effective strategy identified as that with the highest NHB [41]. This is mathematically equivalent to the ICER approach. The CHEERS checklist is included in S1 Checklist [49].

#### Budget Impact

In order to account for the effect of implementing 6-months of BPaLM on the national TB program budget in Moldova, we tracked the subset of aforementioned cost outcomes borne by the TB program. We organized these costs under the following categories: drugs, laboratory tests, routine inpatient and outpatient care, and non-routine inpatient and outpatient care (i.e., care stemming from the treatment of grade 4-5 SAEs, for adjustment of a regimen following the detection of resistance on DST, or for LTFU tracing). The estimated budget impact was calculated for each year over a five year period, scaled to the annual number of case notifications of RR-TB in Moldova.

### Statistical analysis

We estimated results via individual-level microsimulation, with lifetime outcomes for each of 10,000 individuals simulated for each of the diagnostic and treatment strategies described above.

#### Sensitivity analyses

Probabilistic Sensitivity Analysis (PSA) was conducted to account for uncertainty by constructing distributions for model input parameters (S1 Table). In a second-order Monte Carlo simulation, we drew 1,000 parameters sets from the distributions. For each parameter set, the 10,000 individuals were simulated through each strategy, and a set of results was calculated. Finally, point estimates for each outcome were calculated as the mean of these 1,000 second-order simulations, and 95% uncertainty intervals (UIs) were constructed using the 2.5^th^ and 97.5^th^ centiles [50]. Point estimates and 95% UIs were also calculated for the differences between leading 6 month BPaLM-based and SOC-based strategies, and p-values were constructed from the empirical cumulative distribution function of those differences. Further detail is provided in the S1 Appendix.

Some important model parameters have substantial uncertainty. We performed one-way sensitivity analyses on two of these key inputs to understand the relationship with study outcomes. First, we varied the main effect estimate for cure across the uniform distribution (1.00, 2.14). Next, we varied the prevalence of fluoroquinolone resistance among individuals with diagnosed RR-TB across the uniform distribution (0%, 40%) to aid the generalization of results to settings with a different prevalence of fluoroquinolone resistance.

#### Validation

We validated the modeled end-of-treatment (EOT) outcomes to estimates reported to WHO over the period 2010-2019. We also validated the composite of unfavorable outcome at 72 weeks against the findings of TB-PRACTECAL [8]. Further detail is provided in the S1 Appendix.

#### Software

The simulation was conducted in TreeAge Pro Healthcare 2023 [51] and figures were made in R [52] using several packages [53–61]. TreeAge and R code files are available in a repository.[62]

## RESULTS

### Health effects, costs, and cost-effectiveness

Health effects, costs, and cost-effectiveness results for all strategies are presented in Table 3 and Fig 1. Among the 6-month BPaLM strategies, the highest health benefits were achieved by Strategy (1) (BPaLC if Mfx stopped, second-line DST upfront, then repeated at 4 monthly intervals), with undiscounted QALYs of 14.75 (95% UI: [12.76, 16.54]). The two standard of care strategies (Strategies (7) and (8)) both produced slightly more QALYs than Strategy (1), with less than 0.01 undiscounted QALYs between them on average. The Life Years (unadjusted for health-related quality of life) obtained under each strategy are displayed in S3 Table.

**Fig 1.**
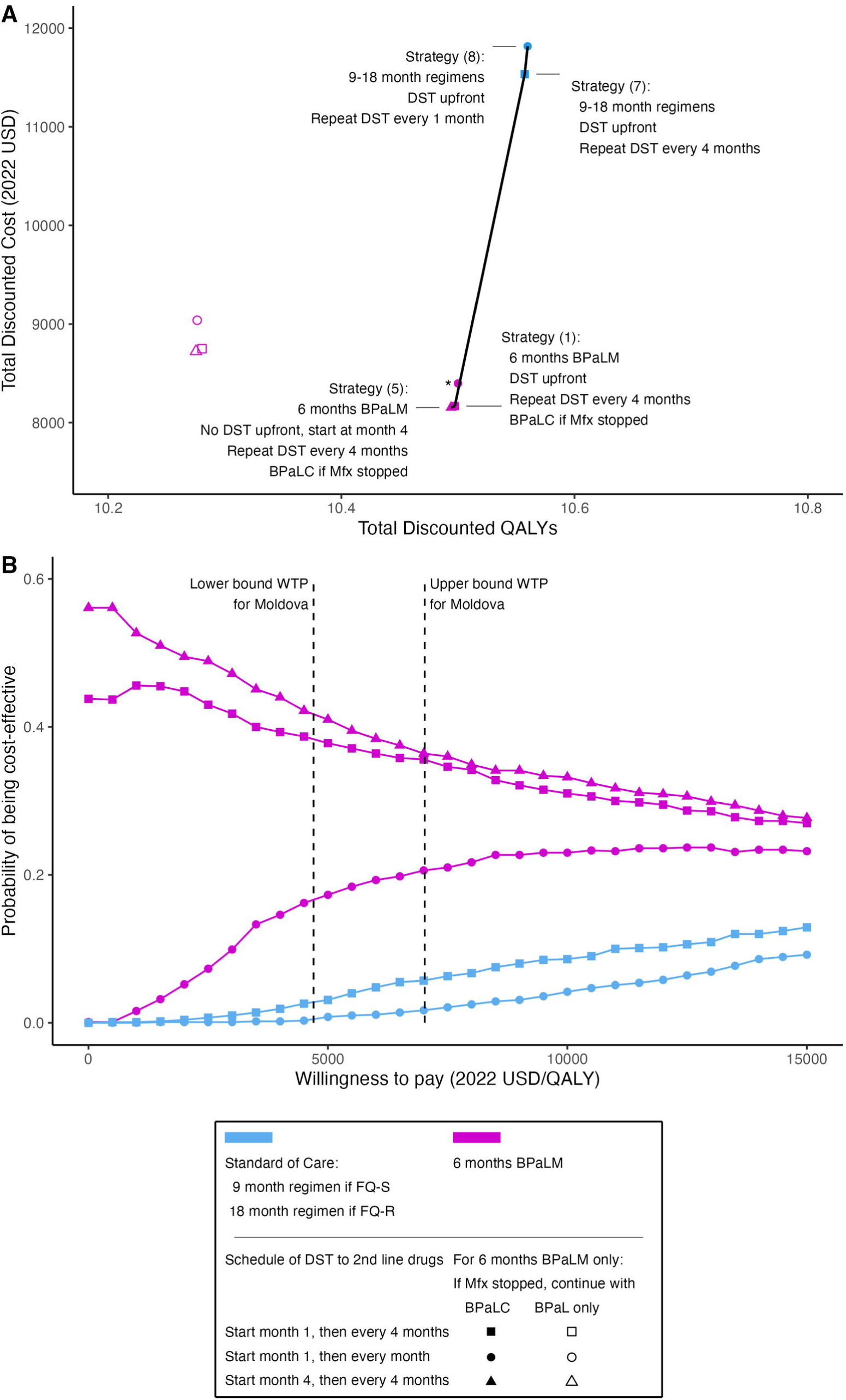
6 months of BPaLM is cost-effective in treating RR-TB. (A) The cost-effectiveness plane shows point estimates for the discounted total costs and discounted QALYs under each modeled strategy. These are calculated as the mean of all simulation runs (1,000 second order Monte Carlo simulations, each with 10,000 individual patient simulations). The efficient frontier (black lines) connects the non-dominated strategies based on point estimates. (B) The cost-effectiveness acceptability curve displays the probability that each modeled strategy is the most cost-effective strategy at different levels of WTP. This probability is calculated as the proportion of 1,000 second-order Monte Carlo simulations where the respective strategy was optimal, given the value for WTP. Strategies were excluded if they were not cost-effective in any of the simulations (these were strategies 3, 4, and 6, where BPaL only was used if Mfx had to be stopped under a BPaLM regimen). Vertical dashed lines mark the lower and upper bounds of the WTP thresholds for Moldova. *Strategy (2) (6 months BPaLM, BPaLC if Mfx discontinued, DST upfront then every 1 month) was very close to the efficient frontier but was dominated by extended dominance based on point estimates. BPaL, bedaquiline, pretomanid, linezolid; BPaLC, bedaquiline, pretomanid, linezolid, clofazimine; BPaLM, bedaquiline, pretomanid, linezolid, moxifloxacin; DST, drug susceptibility testing; FQ-R, fluoroquinolone-resistant; FQ-S, fluoroquinolone-susceptible; Mfx, moxifloxacin; QALY, quality-adjusted life year; USD, United States dollars; WTP, willingness-to-pay.

**Table 3.** Costs, health impacts, and cost-effectiveness of RR-TB treatment strategies.

| STRATEGY DESCRIPTION |  |  |  | COST |  | HEALTH IMPACT |  | COST-EFFECTIVENESS |  |  |  |  |
| --- | --- | --- | --- | --- | --- | --- | --- | --- | --- | --- | --- | --- |
| Strategy Name | Alternative regimen if Mfx stopped (2022 Guidelines only) | 2<sup>nd</sup> line DST at treatment initiation | Routine frequency of subsequent 2<sup>nd</sup> line DST | Undiscounted Total Cost (2022 USD) | Discounted Total Cost (2022 USD) | Undiscounted QALYs | Discounted QALYs | Incremental Discounted Total Cost (2022 USD) | Incremental Discounted QALYs | Incremental Cost-effectiveness Ratio | NHB, lower bound WTP (QALYs) | NHB, upper bound WTP (QALYs) |
| (5) 6 months BPaLM | BPaLC | No | Every 4 months | 8,412<br>(6,469, 10,974) | 8,153<br>(6,279, 10,592) | 14.745<br>(12.72, 16.55) | 10.494<br>(9.26, 11.5) | (comparator) | (comparator) | (comparator) | 8.759<br>(7.42, 9.87) | 9.333<br>(8.07, 10.39) |
| **(1) 6 months BPaLM** | **BPaLC** | **Yes** | **Every 4 months** | **8,424**<br>**(6,469, 10,991)** | **8,167**<br>**(6,299, 10,629)** | **14.750**<br>**(12.76, 16.54)** | **10.497**<br>**(9.28, 11.52)** | **14**<br>**(-170, 205)**<br>**p=0.876** | **0.0032**<br>**(-0.16, 0.19)**<br>**p=0.996** | **4,375** | **8.759**<br>**(7.46, 9.90)** | **9.334**<br>**(8.09, 10.43)** |
| (2) 6 months BPaLM | BPaLC | Yes | Every 1 month | 8,663<br>(6,713, 11,219) | 8,398<br>(6,518, 10,767) | 14.753<br>(12.74, 16.49) | 10.500<br>(9.28, 11.50) | -- | -- | Dominated* | 8.713<br>(7.40, 9.83) | 9.304<br>(8.05, 10.38) |
| (6) 6 months BPaLM | BPaL only | No | Every 4 months | 9,059<br>(6,926, 11,876) | 8,723<br>(6,707, 11,394) | 14.405<br>(12.44, 16.10) | 10.275<br>(9.07, 11.24) | -- | -- | Dominated | 8.419<br>(7.10, 9.51) | 9.033<br>(7.78, 10.07) |
| (3) 6 months BPaLM | BPaL only | Yes | Every 4 months | 9,085<br>(6,935, 11,925) | 8,750<br>(6,703, 11,436) | 14.414<br>(12.42, 16.17) | 10.280<br>(9.10, 11.29) | -- | -- | Dominated | 8.419<br>(7.07, 9.55) | 9.034<br>(7.80, 10.09) |
| (4) 6 months BPaLM | BPaL only | Yes | Every 1 month | 9,391<br>(7,201, 12,126) | 9,039<br>(6,950, 11,635) | 14.408<br>(12.42, 16.16) | 10.276<br>(9.08, 11.28) | -- | -- | Dominated | 8.353<br>(7.04, 9.50) | 8.989<br>(7.71, 10.08) |
| (7) Standard of care | N/A | Yes | Every 4 months | 11,936<br>(9,403, 15,102) | 11,534<br>(9,088, 14,619) | 14.832<br>(13.00, 16.54) | 10.557<br>(9.46, 11.54) | 3,366<br>(1,465, 5,742)<br>p=<0.001 | 0.0600<br>(-0.32, 0.49)<br>p=0.79 | 56,100 | 8.103<br>(6.88, 9.19) | 8.915<br>(7.77, 9.92) |
| (8) Standard of care | N/A | Yes | Every 1 month | 12,232<br>(9,727, 15,435) | 11,816<br>(9,395, 14,912) | 14.836<br>(12.98, 16.56) | 10.560<br>(9.46, 11.55) | 282<br>(24, 550)<br>p=0.028 | 0.0025<br>(-0.16, 0.18)<br>p=0.996 | 112,800 | 8.045<br>(6.83, 9.10) | 8.877<br>(7.73, 9.87) |

Strategies listed in order of increasing discounted total cost. Lower bound willingness-to-pay = 4,700 USD/QALY; upper bound willingness-to-pay = 7,021 USD/QALY. Dominated strategies are those that were both more costly and resulted in poorer health than at least one other strategy, based on point estimates. For nondominated strategies (i.e., strategy numbers 1, 5, and 7), the cheapest (Strategy 5) is listed as the comparator. For strategies 1, 7 and 8, incremental discounted total cost and incremental discounted QALYs were calculated relative to the next cheapest, nondominated strategy. The most cost-effective strategy is highlighted in **bold** text. Mean values are shown with accompanying 95% UIs in parentheses.
\*Strategy (2) was dominated by extended dominance; ICER = 77,000 USD/QALY

Strategy (5) (6-months BPaLM, second-line DST at 4 months and then every 4 months, BPaLC if Mfx stopped) had the lowest undiscounted lifetime total costs ($8412, 95% UI: [6469, 10991]), followed by Strategy (1) and Strategy (2) (Table 3).

Compared to 6-month BPaLM-based strategies where BPaLC was used if Mfx had to be stopped, strategies continuing only the three-drug regimen BPaL (Strategies (3), (4), and (6)) resulted in worse overall health *and* additional lifetime total costs. The frequency of second-line DST did not lead to large differences in health or cost outcomes (Fig 1).

We compared cost-effectiveness ratios (ICERs) to current cost-effectiveness criteria for Moldova, with the willingness-to-pay for health improvements assumed to fall between $4700 and $7021 per QALY gained. According to this approach Strategy (1) (6-months BPaLM, DST upfront then every 4 months, BPaLC if Mfx stopped) was the most cost-effective strategy with an ICER of $4375 per QALY.

Strategy (7) was potentially cost-effective, but only with a willingness to pay over $56,100 per additional QALY, far higher than the upper bound threshold. In Fig 1B, we show the probability that each strategy is the most cost-effective for given cost-effectiveness thresholds.

Strategies (1), (2), and (5) had the highest probabilities of being cost-effective. All three are 6-month BPaLM strategies where BPaLC was continued for those stopping Mfx, and differ only based on the schedule of routine DST. Taken together, the probability that one of these strategies would be most cost-effective was at least 93% across the range of cost-effectiveness thresholds for Moldova.

For simplicity, we henceforth make comparisons between the leading (i.e., most cost-effective based on point estimates) 6-month BPaLM-based and standard of care-based strategies: Strategy (1) and Strategy (7), respectively. The category-specific costs for these strategies are shown in Fig 2, and the incremental cost-effectiveness for this one-to-one comparison in S7 Fig.

**Fig 2.**
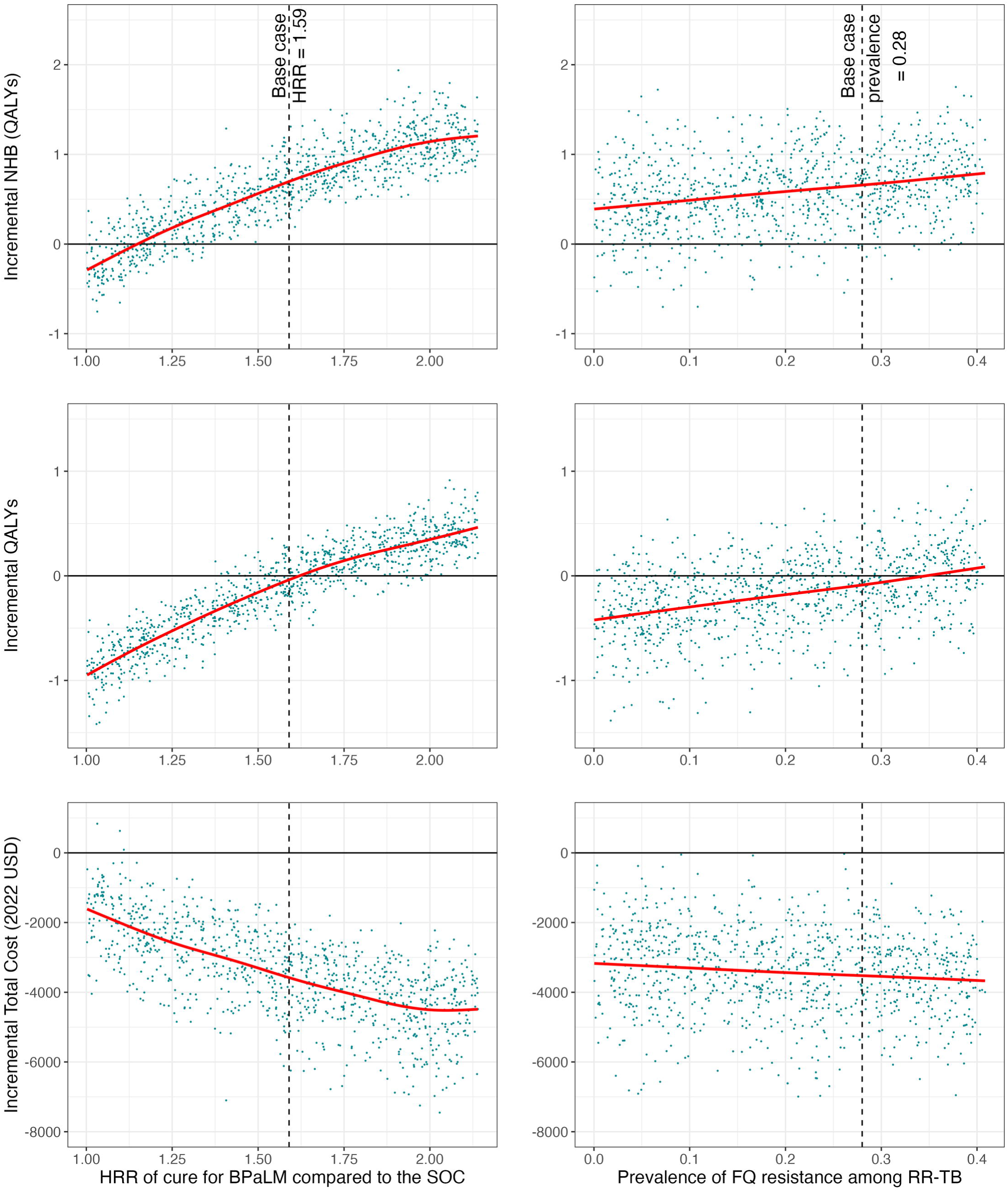
Lifetime costs for 6 months BPaLM and Standard of Care strategies by category. Undiscounted lifetime costs per individual for Strategy (1) (6 months BPaLM, DST upfront, repeat DST every 4 months, BPaLC if Mfx stopped) as compared to Strategy (7) (standard of care 9-18 month regimens based on results of upfront DST, repeat DST every 4 months). The bars show the mean model outcomes for each cost category, with error bars representing 95% UIs. BPaLC – bedaquiline, pretomanid, linezolid, clofazimine; BPaLM – bedaquiline, pretomanid, linezolid, moxifloxacin; DST – drug susceptibility testing; UI – uncertainty interval; USD – United States Dollars

Compared to the standard of care (Strategy (7)), the incremental NHB of 6 months BPaLM (Strategy (1)) was 0.656 QALYs; (95% UI [-0.091, 1.383] p=0.082) at the lower bound WTP and 0.419 QALYs; (95% UI [-0.206, 0.994] p=0.166) at the upper bound WTP.

### Drug resistance

When counting time with resistance across the entire cohort, compared to Strategy (7), Strategy (1) was associated with a non-significant change in the mean duration with any RR-TB of −1.10 months; (95% UI [-4.07, 2.28] p=0.486) (Fig 3, S3 Table). Strategy (1) increased the mean duration with resistance to pretomanid by 0.55 months; (95% UI [0.20, 1.05] p<0.001) and delamanid by 0.54 months; (95% UI [0.18, 1.04] p=0.002) (Fig 3, S3 Table).

**Fig 3.**
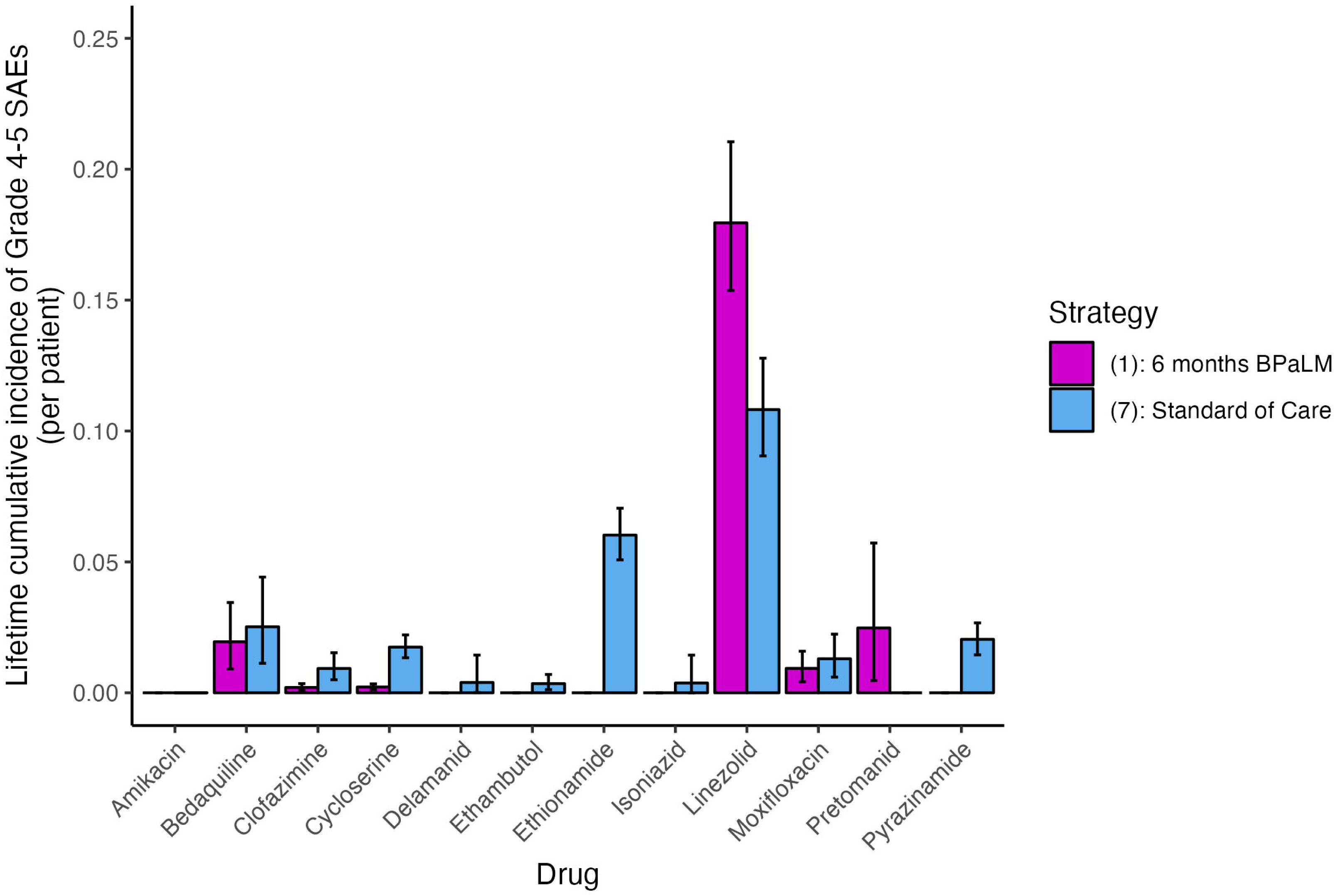
Effect of 6 months BPaLM on duration of resistance to key anti-TB drugs. Results are shown for Strategy (1) (6 months BPaLM, DST upfront, repeat DST every 4 months, BPaLC if Mfx stopped) as compared to Strategy (7) (standard of care 9-18 month regimens based on results of upfront DST, repeat DST every 4 months). For each drug, two estimates are provided: counting time with resistance at any point until the individual is truly cured (dark green), and counting time with resistance only while an individual has TB disease but is not being treated (light green). Both estimates are provided per individual, averaged over the same denominator of the entire cohort initiating treatment. 95% UIs are shown by the accompanying error bars. BPaLC – bedaquiline, pretomanid, linezolid, clofazimine; BPaLM – bedaquiline, pretomanid, linezolid, moxifloxacin; DST – drug susceptibility testing; Mfx, moxifloxacin; SOC – standard of care; TB – Tuberculosis; UI – Uncertainty Interval

In contrast, Strategy (1) decreased the duration with resistance for several drugs: The mean change was - 2.21 months for moxifloxacin (95% UI [-3.39, −1.02] p<0.001), −2.28 months for pyrazinamide (95% UI [-4.02, −0.52] p=0.016), −1.31 months for clofazimine (95% UI [-1.94, −0.80] p<0.001), −0.92 months for bedaquiline (95% UI [-1.48, −0.49] p<0.001), −0.95 months for cycloserine (95% UI [-1.38, −0.62] p<0.001), and −0.40 months for amikacin (95% UI [-0.79, −0.06] p=0.022) (Fig 3, S3 Table). When measuring time with resistance only among those with active, untreated RR-TB, or when measuring lifetime cumulative incidence of resistance, the findings revealed a similar picture (Fig 3, S3 Table).

### Secondary outcomes

Under Strategy (7), the mean number of grade 4-5 SAEs ever experienced per individual was 0.265 (95% UI: 0.233, 0.300). Strategy (1) resulted in a mean number of grade 4-5 SAEs of 0.237 (95% UI [0.197, 0.284]), conferring a decrease of 0.028 grade 4-5 SAEs per person (95% UI [-0.012, 0.063] p=0.17) over the course of treatment. Fig 4 displays the proportion ever experiencing a grade 4-5 SAE to each drug; the point estimates were lower for Strategy (1) than for Strategy (7) for all drugs except linezolid and pretomanid.

**Fig 4.**
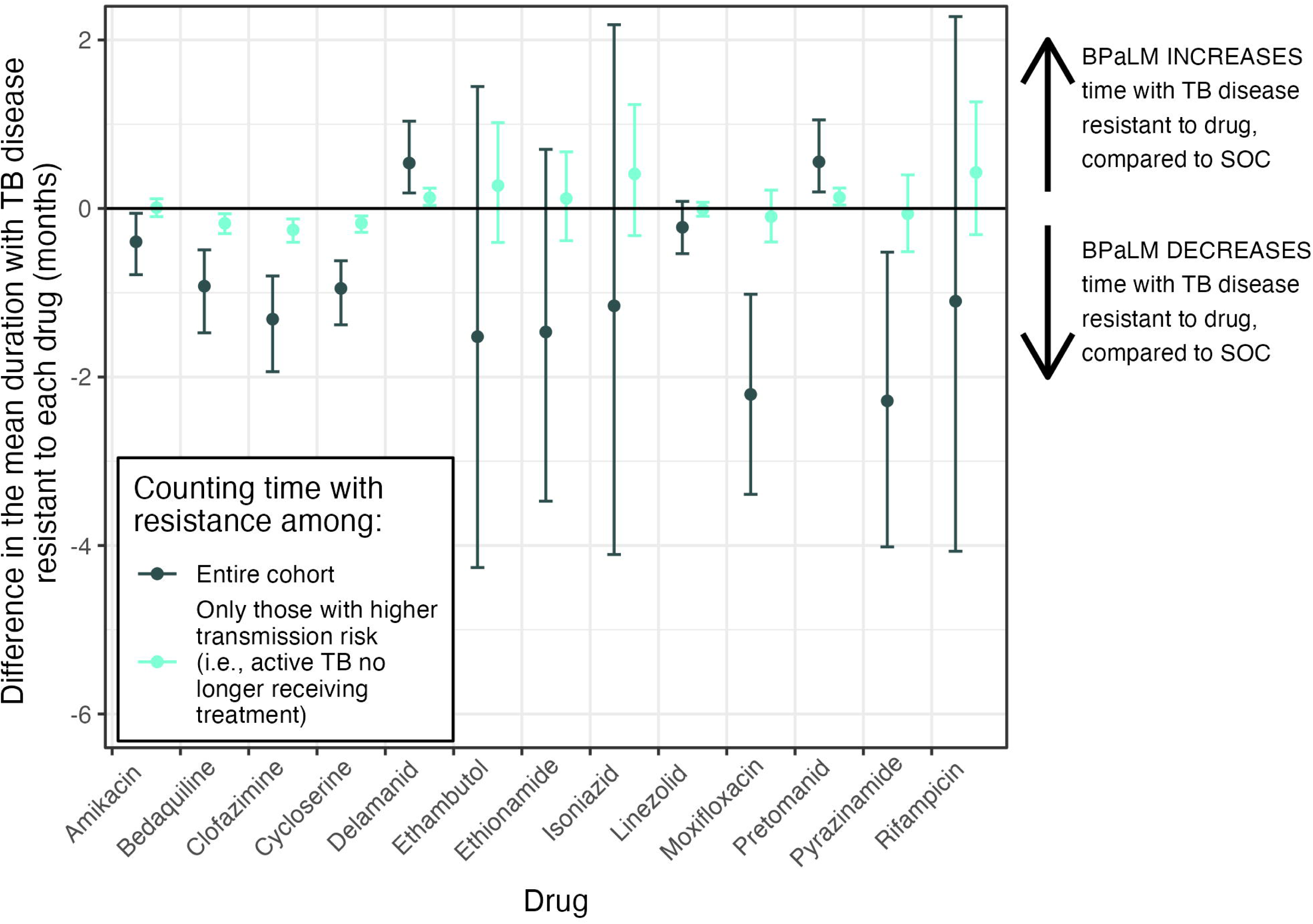
Cumulative incidence of grade 4-5 Severe Adverse Events under 6 months BPaLM and Standard of Care. The mean cumulative incidence of Grade 4-5 Severe Adverse Events ever experienced to each of 12 anti-TB drugs is shown for Strategy (1) (6 months BPaLM, DST upfront, repeat DST every 4 months, BPaLC if Mfx stopped) as compared to Strategy (7) (standard of care 9-18 month regimens based on results of upfront DST, repeat DST every 4 months). Estimates are provided per individual, averaged over the entire cohort initiating treatment. The mean estimate is shown by the bar, with 95% UIs represented as error bars. BPaLM – bedaquiline, pretomanid, linezolid, moxifloxacin; SAE – grade 4-5 Severe Adverse Event; TB – Tuberculosis; UI – Uncertainty Interval

When health benefits were measured using life years unadjusted for health-related quality of life, strategy (7) again conferred a slightly higher life expectancy than strategy (1) on expectation. Also consistent with the primary QALY-based outcomes, the lowest life expectancy was estimated for Strategies (3), (4), and (6) (BPaLM-based strategies where BPaL was continued in the event of Mfx being stopped). (S4 Table).

For the shorter-term endpoints of 6 months, 12 months, and 17 months (i.e., 72 weeks) from treatment initiation, we found that Strategy (1) resulted in a reduction in the composite unfavorable outcome compared to Strategy (7). The reduction was not significant when using the TB-PRACTECAL aligned definitions for unfavorable outcomes, but was significant and larger in magnitude when using WHO-based definitions (S5 Table, S9 Fig).

Compared to Strategy (7), Strategy (1) would be expected to save Moldova’s national TB program budget $7.1 million (95% UI: [1.3 million, 15.4 million] p=0.002) over the five year period from implementation (S6 Table).

### Sensitivity Analyses

Fig 5 shows how cost-effectiveness results change for different values of the hazard rate ratio (HRR) of cure, and the initial prevalence of fluoroquinolone resistance, for Strategy (1) as compared to Strategy (7). In these results, Strategy (1) was estimated to be cost-effective (i.e., had a positive Net Health Benefit) compared to Strategy (7) across the range of values used for these parameters. Similarly, total costs were lower for Strategy (1) compared to Strategy (7) across the range of values assessed. Health outcomes were sensitive to the value of the HRR for cure for the BPaLM regimen as compared to standard of care regimens. For low values of the HRR (HRR = 1), Strategy (1) was estimated to lead to a mean 0.90 reduction in QALYs. For high values (HRR = 2), Strategy (1) would lead to a mean 0.35 gain in QALYs. All data files containing these results are available in a repository.[62]

**Fig 5.**
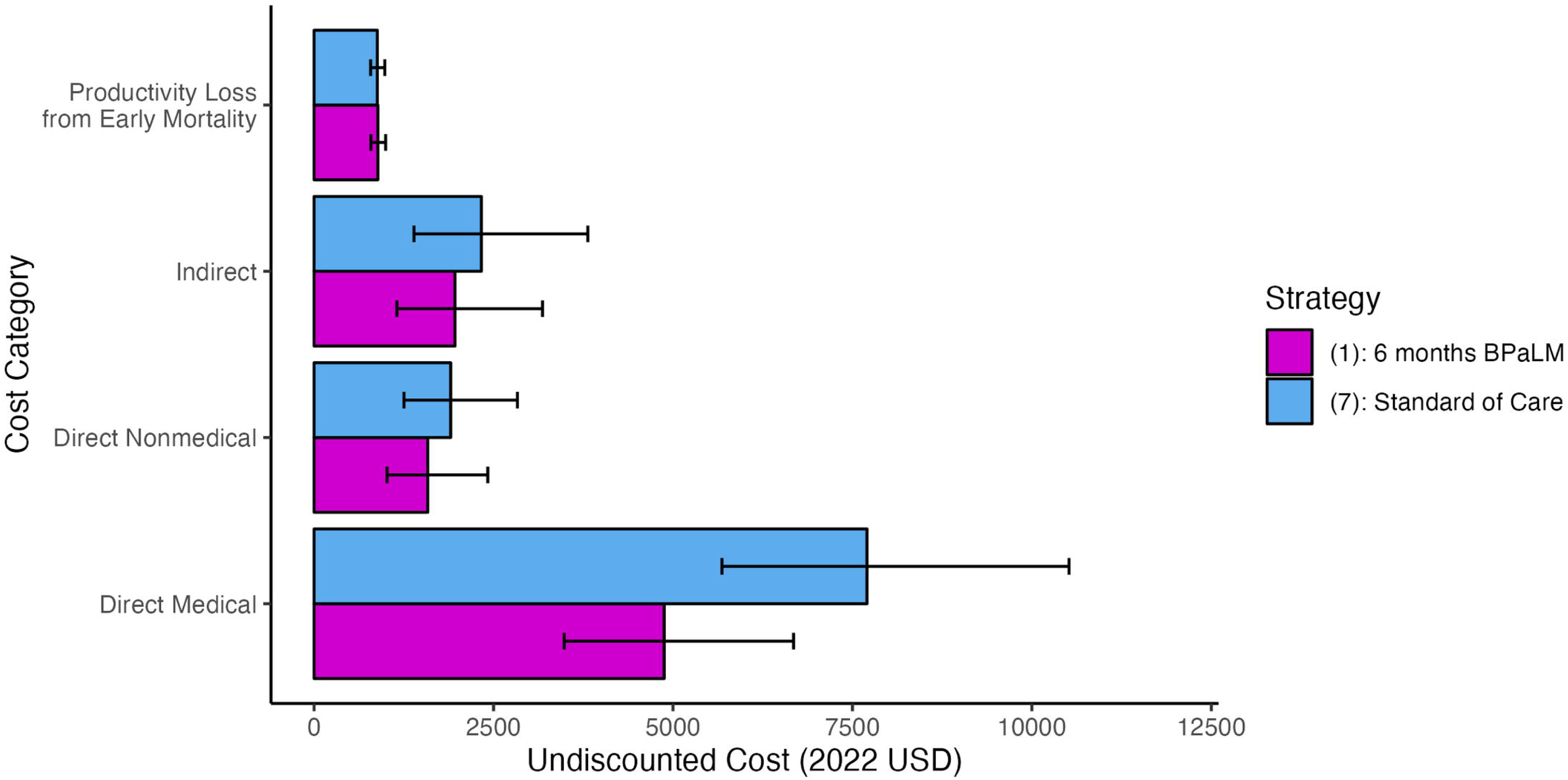
Sensitivity analyses varying relative effectiveness of BPaLM and cohort prevalence of fluoroquinolone resistance. One-way sensitivity analyses exploring the implications of key model parameters, in terms of their effect on the incremental benefits and costs of Strategy (1) (6 months BPaLM, DST upfront, repeat DST every 4 months, BPaLC if Mfx stopped) as compared to Strategy (7) (standard of care 9-18 month regimens based on results of upfront DST, repeat DST every 4 months). We chose to compare these two strategies as they were the best-performing BPaLM-based and standard of care-based strategies, respectively. In the left column, the HRR of cure for the BPaLM regimen compared to the standard of care is varied. In the right column, we vary the starting prevalence of fluoroquinolone resistance in the cohort (i.e., among all RR-TB). Each of the parameters is varied deterministically in the respective sensitivity analysis, with all other model parameters drawn as in the probabilistic sensitivity analysis. The outcomes quantified on the y-axis for each row of plots are (top to bottom): incremental NHB (calculated using discounted Total Costs and discounted QALYs at the lower bound WTP), incremental QALYs (undiscounted), and incremental Total Costs (undiscounted). The difference between the modeled outcomes under BPaLM and the standard of care is shown for 1,000 model runs, each an average of 10,000 individual patient simulations. The red line shows the trend as represented by regression of the y-axis variable on the x-axis variable, using a generalized additive model with cubic spline to obtain a restricted maximum likelihood within ggplot2.[58] The vertical dashed lines mark the base case assumptions for the mean of each of these model parameters. BPaLM – bedaquiline, pretomanid, linezolid, moxifloxacin; FQ – Fluoroquinolone; HRR – Hazard Rate Ratio; NHB – Net Health Benefit QALY – Quality-adjusted Life Year; RR-TB – Rifampicin-resistant tuberculosis

## DISCUSSION

In this study we assessed the potential health impact and cost effectiveness of a 6-month BPaLM regimen for treating RR-TB in a setting with a high prevalence of drug resistance. Compared to a strategy using 9-18 month regimens based on the 2020 WHO treatment guidelines for drug-resistant TB, we found the 6-month BPaLM regimen would be cost-effective across a range of WTP thresholds, with substantial reductions in the duration and cost of treatment, but little expected change in health outcomes. Though there is considerable overlap between some of the 6-month BPaLM implementation scenarios, there is a clear lead for strategies where clofazimine is used to “top up” the regimen if moxifloxacin must be discontinued because of a grade 4-5 SAE or resistant DST result, compared to continuing on the three-drug BPaL regimen alone. Holding the drug regimen constant, the frequency of second-line DST (to fluoroquinolones and injectables using MGIT) did not result in substantial differences to health or cost outcomes.

Though our analysis was principally concerned with health outcomes over the lifetime horizon, our findings also show that 6 months of BPaLM reduces the risk of unfavorable outcomes over shorter time horizons, in line with the direction of effect observed at 72 weeks in the TB-PRACTECAL randomized controlled trial [8]. On cost-effectiveness specifically, our findings for Moldova are in line with a previous economic evaluation for populations across South Africa, Belarus, and Uzbekistan [28]. Belarus also has a high proportion of RR-TB among individuals newly diagnosed with TB [2], but we do not know whether the joint distribution of resistance to other important drugs would differ between Belarus and Moldova. Although South Africa and Uzbekistan have a lower prevalence of resistance to many drugs, we found that 6 months of BPaLM remained cost-effective when the proportion of RR-TB patients with FQ-R was varied across the wide range of 0-40% (compared to Moldova at 28%). Our analysis builds on the aforementioned cost-effectiveness analysis by explicitly modelling the acquisition of drug resistance, with the initial cohort resistance profile informed by genetic sequencing data from Moldova. We also investigated the potential effects of a larger number of policy implementation scenarios, including the frequency of DST, and whether patients having to stop Mfx under BPaLM should continue on BPaL alone or continue on an alternative four-drug regimen.

When modeling the effectiveness estimate for BPaLM as compared to the standard of care, we assumed that the treatment effect for true cure in the model was approximated by the treatment effect for sputum culture conversion from the TB-PRACTECAL trial [8]. Although the trial measured clinical outcomes, its primary composite outcome measure combined treatment failure, discontinuation, LTFU, death and recurrence, outcomes that are important to distinguish to calculate long-term health outcomes. The numbers of individuals experiencing each of the long-term outcomes of greatest clinical interest were very small. Even if the effect on true cure is not the same as on culture conversion, we found that 6-months BPaLM remained the cost-effective strategy when the HRR (point estimate: 1.59) was varied over a wide range.

While both regimens perform best at lower levels of resistance, sensitivity analyses showed that 6 months of BPaLM may result in a reduction in total QALYs as compared to the standard of care at lower levels of initial FQ-R, or if the BPaLM regimen has lower comparative effectiveness than estimated in the TB-PRACTECAL trial, even while it provides overall value for money. Although policymakers may be uncomfortable adopting interventions that reduce health benefit on expectation, this difference was not statistically significant. Adopting the new regimen would bring substantial benefits in the form of reduced regimen duration, and freeing up funding to spend on other health interventions.

In this analysis we found that 6 months of BPaLM improved or resulted in no change to the duration of disease with resistant strains of *M. tuberculosis* as well as the cumulative incidence of resistance for all anti-TB drugs investigated, except pretomanid and delamanid. Both the duration and cumulative incidence measures were influenced by the starting profile of resistance as informed by the WGS data, the rate of acquisition of new resistance to each drug under each modeled drug regimen, and the monthly rate of cure. Changes in the rate of acquisition of resistance are important for individuals undergoing treatment today (some of the effects of this are captured in the QALYs estimated under each strategy) but preventing new second-line resistance is also important for the health outcomes of those living with RR-TB in the future.

This analysis had several limitations. The Moldovan genomic data used to characterize the resistance profile in the modeled population were from culture positive sputum specimens in 2018-19, and so may not accurately describe current resistance patterns in Moldova or resistance elsewhere, although we hope the sensitivity analysis on the prevalence of FQ-R aids in the generalization of findings. Because the publicly-available WGS dataset excluded samples with mixed strains of *M. tuberculosis* (17.4%), it is possible that our findings do not adequately address this subpopulation with mixed infections, although we note that all the remaining model parameters reflect the real-world health outcomes and costs of a mix of mono- and mixed-strain infections. Furthermore, we assumed that that the true resistance profile was perfectly predicted by the presence or absence of mutations conferring resistance in this data: While the sensitivity and specificity of genomic sequencing is very high for detecting resistance in rifampicin, isoniazid, and ethambutol, the performance is less favorable for moxifloxacin, amikacin, and ethionamide [63].

There are also limitations pertaining to the simulation of health and cost outcomes. The hazard rate ratio for cure was based on the outcome of sputum culture conversion from TB-PRACTECAL; while culture conversion is indeed a prognostic marker in TB [64], it is not a perfect substitute to quantify the rate of true cure, which is unobservable. Further, real-world outcomes with 6-months of BPaLM are likely to be less favorable than in the high-fidelity environment of a randomized controlled trial—for example, there may have been a higher frequency of follow-up in the trial—and the status quo may differ between settings. We did not explicitly model the differences in adherence that may exist between regimens, and we made the simplifying assumption that increasing the number of effective drugs increases the monthly rate of cure and reduces the rate of acquiring resistance. This was based on a previously applied approach [7] and is likely to hold qualitatively, but we did not account for the all the differences that may exist between specific drugs, and the interactions between them. For example, the effectiveness of BPaLC vs. BPaL may not be the same as the effectiveness of BPaLM vs. BPaL, yet—SAEs aside—the modeling approach was agnostic to this, conditional on the number of “effective” drugs in the regimen. For parsimony, we did not explicitly model changes in smear status. For individuals no longer receiving treatment, we adopted a mortality rate estimate for smear-positive TB, which may overestimate mortality specifically for those who appear to have completed treatment successfully but not truly cured; this would likely bias the results against shorter, 6-month BPaLM strategies. While the relationship between HIV and RR-TB treatment outcomes is neither straight-forward nor consistent [38,65], we did not model HIV status at the individual level and as such we were unable to comment specifically on health outcomes for those with TB-HIV coinfection. Although the probability of a grade 4-5 SAE was modelled separately for each drug, we did not incorporate variation in the duration and consequences of each type of SAE. Finally, we did not account for the secondary effects resulting from onward transmission of RR-TB, and our results may therefore not capture the full cost-effectiveness implications of each modelled strategy.

To account for this explicitly, it would be necessary to model the transmission dynamics of *M. tuberculosis*. Instead, we estimated the cumulative incidence and duration of resistance as surrogates for the long-term health outcomes they may affect, insofar as lower incidence and fewer months of resistant disease might each result in less transmission of resistant strains.

This study was conducted in the setting of Moldova, a country with a high proportion of RR-TB with resistance to second-line drugs. By conducting sensitivity analysis on the proportion with FQ-R, we aimed to aid the generalization of findings to other settings. Many of the health-related model parameters are also generalizable beyond Moldova: TB outcomes under the standard of care were informed by multi-national meta-analyses, and the estimate for comparative effectiveness was from a multi-national trial (S1 Table). However, many of the cost parameters were from Moldova and Georgia (GDP per capita of $5,563 and $6,628 in 2022, respectively) [66], and so there are likely limitations in the generalization of incremental costs of 6 months BPaLM compared to the standard of care, especially to countries with very different income levels.

To optimize clinical care for RR-TB, decision makers must take account of important health and economic effects for affected individuals as well as society at large. In this study, we estimated favorable cost-effectiveness for the 6-month BPaLM regimen in settings with a high burden of drug resistance, conditional on BPaLC being used in the event of moxifloxacin being contraindicated, rather than BPaL alone. The schedule of second-line DST did not appear to affect health outcomes or costs to a great degree across the finite number of DST schedules we explored, and further analyses may be warranted to explore the optimal testing frequency in Moldova and other settings—especially where second-line DST capacity is limited or unavailable [67]—and to explore additional technologies beyond MGIT for identifying resistance to fluoroquinolones and injectables. The forthcoming results of the endTB trial [9–13] will expand the evidence base for shorter regimens, and while the trial investigated 9-as opposed to the 6-month regimens investigated in this study, this still represents a substantial shortening compared to many standard of care regimens. The growing body of both empirical and modeling literature may also highlight the elements of treating RR-TB—including the choice of drugs, duration of regimen, and frequency and modality of DST—which overall provide the best treatment strategy, for each patient’s specific needs. Clinical and health policy decisions alike would be enhanced by continued collective efforts to strengthen the evidence base in the ways most likely to optimize care, with sufficient numbers of patients to quantify long-term health outcomes across multiple settings.

## Supporting information

Supplemental Checklist 1

## Data Availability

All data produced in the present study are either contained in the manuscript or are available upon reasonable request to the authors.

https://www.ncbi.nlm.nih.gov/bioproject/PRJNA736718

https://github.com/lyndonpjames/BPaLM_Moldova

## Acknowledgments

For helpful feedback on research-in-progress presentations, LPJ would like to thank current and former students, postdocs and faculty affiliated with: the Center for Health Decision Science, the PhD Program in Health Policy, and the Center for AIDS Research, all at Harvard University, Cambridge, MA, USA; the Decision Science Methods Group at Erasmus University, Rotterdam, the Netherlands.

## Author Contributions

Conceptualization: LPJ, NAM, TC, JF, SS, RY, VC, NC, AC

Data curation: LPJ, TC, NAM

Formal Analysis: LPJ, NAM

Funding Acquisition: LPJ, NAM, TC

Investigation: LPJ, NAM, TC, MFF

Methodology: LPJ, NAM, SS, RY

Project Administration: LPJ, NAM

Resources: LPJ, NAM

Software: LPJ, NAM, FK

Supervision: NAM, TC

Validation: LPJ, TC, MFF, NAM, DC, JF, VC

Visualization: LPJ, FK, NAM

Writing – Original Draft: LPJ, NAM

Writing – Review & Editing: LPJ, NAM, MFF, TC, JF, FK, SS, RY, DC, NC, VC, AC

## Abbreviations

BPaL: bedaquiline, pretomanid, linezolid
BPaLC: bedaquiline, pretomanid, linezolid, clofazimine
BPaLM: bedaquiline, pretomanid, linezolid, moxifloxacin
CDF: cumulative distribution function
CI: confidence interval
CrI: credibility interval
CPI: Consumer Price Index
DST: drug susceptibility testing
FQ-R: fluoroquinolone-resistant
FQ-S: fluoroquinolone-susceptible
LJ: Lowenstein-Jensen
MDL: Moldovan Leu
GDP: Gross Domestic Product
GEL: Georgian Lei
HIV: human immunodeficiency virus
LTFU: lost to follow up
LY: Life Year
*M. tb.*: *Mycobacterium tuberculosis*
MDR-TB: multidrug-resistant tuberculosis
NHB: Net Health Benefit
QALY: Quality-adjusted Life Year
RR-TB: rifampicin-resistant tuberculosis
SAE: grade 4-5 Severe Adverse Event
SEM: Standard Error of the Mean
SMR: standardized mortality ratio
TB: tuberculosis
UI: Uncertainty Interval
USD: United States Dollars
WTP: Willingness-to-Pay
XDR-TB: extensively drug-resistant tuberculosis

