## Supplemental Appendix 2 for "Impact and cost-effectiveness of the 6-month BPaLM regimen for rifampicin-resistant tuberculosis: a mathematical modeling analysis"

### Calculated Model Parameters

**Note on terminology:** As in most of the modeling literature, but not always the wider public health literature, we use the term rate and probability/proportion to mean different things. The former is in the sense of an instantaneous rate of change. Unlike a probability/proportion, it can be greater than 1. To convert between the two when the rate is constant over time, we use $P(X) = 1 - e^{-r*t}$, where $P(X)$ is the probability of an event $X$ occuring in a time period of length $t_1$, $r$ is the event rate per unit of time $t_2$, and $t = \frac{t_1}{t_2}$.

There are several model parameters that are not well approximated by the available data. In some cases, we adjusted published estimates to better fit the model's processes. Details for some of these parameter calculations are summarized in Table $\mathbb{S}1$, but here we provide some additional notes on calculations for some parameters.

#### 1. The probability of being correctly identified as not cured (i.e., non-success), conditional on being truly not cured

"True" cure here refers to the elimination of *M. tuberculosis* in the host, such that it is no longer capable of causing disease. This is contrasted with an observed cure, where the patient clinically appears to have successfully completed treatment, but may subsequently have recurrent disease. "TB" here refers to all RR-TB specifically.

This is an unobserved (i.e. latent) process and as such cannot be estimated from empirical observation.

Instead, we use an estimate from Blöndal et al 2012. This paper reported that among 129 patients observed to be successfully treated at the end of their TB regimen, 11 had recurrent disease over a median follow-up time of 98.1 months.

The paper also demonstrates that approximately 90% of the cohort had event-free survival by the end of follow-up (Fig. 4 shows Kaplan-Meier curves separately for: a) non-XDR but MDR-TB, and b) XDR-TB. Though there was varying duration of follow-up, we also apply the 98.1 month median follow-up duration here for calculations.

For simplicity, we make the following assumptions with regard to this study:

- we assume that all recurrences in this paper are relapses returning to treatment, not reinfection
- we assume that *all* patients who are truly cured are correctly identified as such, and thus do not receive a treatment extension
- we assume that there is no self-cure applied to this group

The following diagram represents some of the processes of interest here.

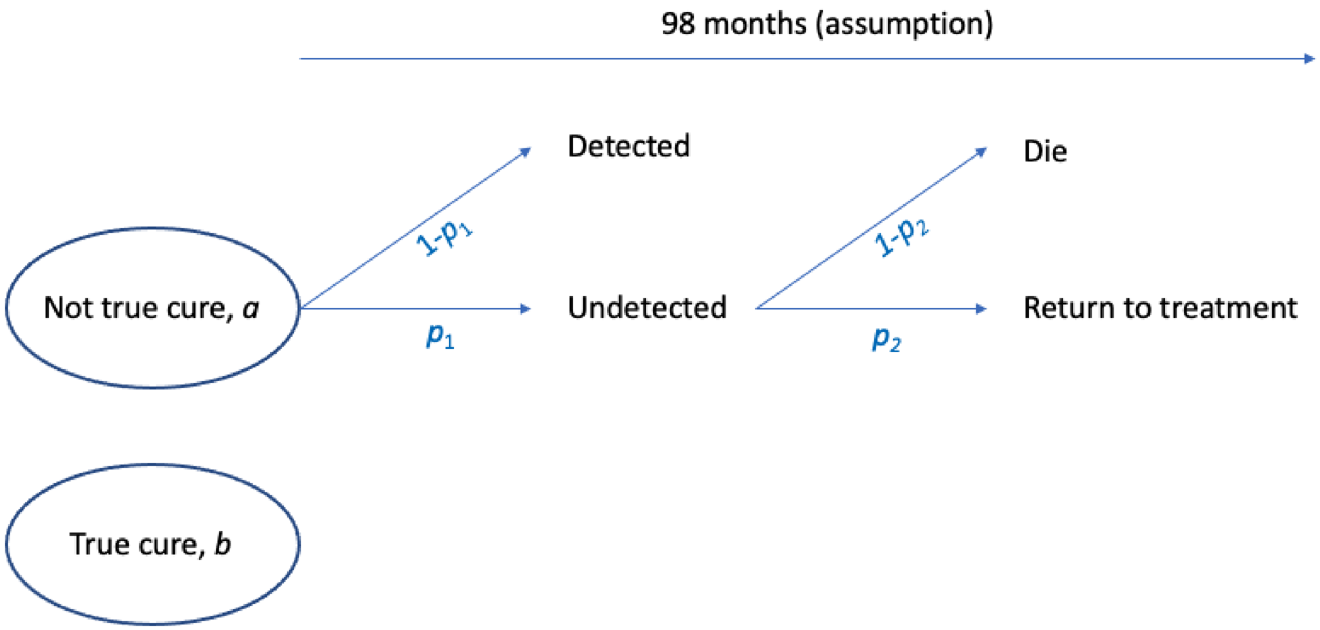


The quantities of interest in the figure are as follows:

- 98 months, the median follow-up time in the study, adopted as the timeframe for this process
- $a$, the proportion who are not truly cured among those who complete a TB treatment regimen,
- $b = 1 - a$ be the proportion who are truly cured among those who complete a treatment regimen,
- $p_1$, the proportion of those not truly cured who are undetected/misclassified. These individuals are observed to be successfully treated, and thus are discharged from care.
- $p_2$, the proportion of those undetected, not truly cured individuals who eventually return to treatment by 98 months.
- $1 - p_2$, the proportion of those undetected, not truly cured individuals who die by 98 months,
- individuals who are not truly cured and are correctly detected receive a treatment extension as described elsewhere,

The estimate from the study can be represented as follows:

Pr(return to treatment|observed successfully treated) $= q = \frac{11}{129} = \frac{ap_1p_2}{ap_1+b}$

Defined for this population, (i.e., those observed to have been successfully treated):

Let $C$ be the event that such an individual has truly been cured.

Let $R$ be the event that such an individual returns to treatment.

Let $D$ be the event that such an individual dies.

We start therefore with

$$P(R) = \frac{11}{129}$$
$$P(R \cup D) \approx 1 - 0.9 = 0.1$$

Because an individual would only be recorded as returning to treatment or dead, not both,

$$P(R \cap D) = 0$$

which implies

$$P(R \cup D) = P(R) + P(D) - P(R \cap D) = P(R) + P(D) + 0$$
$$= 0.1$$

and thus,

$$P(R \cup D) = P(R) + P(D)$$
$$0.1 = \frac{11}{129} + P(D)$$
$$P(D) = 0.1 - \frac{11}{129}$$
$$P(D) = 0.0147$$

Applying LOTP to $P(R)$,

$$P(R) = P(R|C)P(C) + P(R|\bar{C})P(\bar{C})$$

Because no individual who has truly cured will relapse, this simplifies to

$$P(R) = 0 * P(C) + P(R|\bar{C})P(\bar{C})$$
$$P(R) = P(R|\bar{C})P(\bar{C})$$

Let us assume that, by 98 months, almost all (let us say 95%) of individuals who were not truly cured at the end of treatment will have either returned to treatment or died.

Let the monthly rate of relapse be $\mu_R$.

Let the monthly mortality rate of TB on treatment be $\mu_{D\ on\ Rx}$. The one month probability of death while on treatment is 0.004,

$$\begin{aligned}\mu_{D\ on\ Rx} &= -ln(1-0.004)\\ &= 0.004008\end{aligned}$$

To determine $P(D|\bar{C})$,

$$\begin{aligned}P(D|\bar{C}) &= 1-e^{-0.004008*98}\\ &= 0.3248\end{aligned}$$

And so,

$$\begin{aligned}P(D\cup R|\bar{C}) &= 0.95\\ 98(\mu_{D\cup R}) &= -ln(1-0.95)\\ 98(\mu_{D\ on\ Rx}+\mu_R) &= -ln(1-0.95)\\ 98(\mu_{D\ on\ Rx}+\mu_R) &= 3\\ \mu_{D\ on\ Rx}+\mu_R &= \frac{3}{98}\\ 0.004008+\mu_R &= \frac{3}{98}\\ \mu_R &= 0.0266\end{aligned}$$

assuming a constant relapse rate over the course of 98 months.

Let us also use this estimate to inform the proportion of this cohort not truly cured. Recall

$$\begin{aligned}P(R) &= P(R|\bar{C})P(\bar{C})\\ \frac{11}{129} &= P(R|\bar{C})P(\bar{C})\end{aligned}$$

Because of the competing events of death and relapse in this problem where only one is recorded, I simulated a Markov model over 98 monthly cycles using the rates calculated above. After 98 months, 82.5% of the not truly cured had returned to treatment, and 12.5% had died without returning to treatment. As such,

$$\begin{aligned}\frac{11}{129} &= P(R|\bar{C})P(\bar{C})\\ \frac{11}{129} &= 0.825*P(\bar{C})\\ P(\bar{C}) &= 0.103\end{aligned}$$

**So in this population of individuals who had been observed to successfully complete treatment, we estimate that approximately 10.3% were not truly cured of the disease.**

Now let's obtain the estimate for the proportion misclassified.

Referring back to the diagram and associated notation, we can express $P(\bar{C}) = 1 - P(C)$ as

$$1 - P(C) = \frac{ap_1}{ap_1 + b}$$
$$0.103 = \frac{ap_1}{ap_1 + b}$$
$$0.103 = \frac{ap_1}{ap_1 + 1 - a}$$

While $a$ and $b = 1 - a$ will change based on the cohort characteristics, treatment and other factors, we model $p1$ as a variable independent of the overall proportion who truly have disease, i.e. it is a test characteristic, 1-sensitivity of a clinical assessment "test" to detect those with the "disease" of being truly cured.

The proportion with observed success in the study by Blondal et al. 2012 is 129/211 = 0.611 We note that this is approximately the same as the overall proportion with observed success in the Bastos et al. 2017 review of 0.64.

This can be represented as the denominator of the expression above,

$$ap_1 + b = 0.611$$
$$ap_1 + 1 - a = 0.611$$
$$ap_1 - a = 0.611 - 1$$
$$a - ap_1 = 1 - 0.611$$
$$a(1 - p_1) = 1 - 0.611$$
$$a = \frac{1 - 0.611}{1 - p_1}$$
$$a = \frac{0.389}{1 - p_1}$$

This can now be substituted into the equation above to solve for $p_1$.

$$0.103 = \frac{ap_1}{ap_1 + 1 - a}$$
$$0.103 = \frac{\frac{0.389p_1}{1-p_1}}{1 + \frac{0.389p_1}{1-p_1} - \frac{0.389}{1-p_1}}$$
$$0.103 = \frac{0.389p_1}{1 - p_1 + 0.389p_1 - 0.389}$$
$$0.103 = \frac{0.389p_1}{0.611 - 0.611p_1}$$
$$0.103(0.611 - 0.611p_1) = 0.389p_1$$
$$0.0629 - 0.0629p_1 = 0.389p_1$$
$$0.0629 = 0.452p_1$$
$$p_1 = 0.139$$

**We obtain an estimate that 13.9% of patients who are truly not cured are misclassified as cured at the end of treatment (i.e. clinical assessment is 84.1% sensitive in being able to detect those not truly cured).**

#### 2. Calculating different mortality rates for those cured and not cured, while on treatment

There are estimates from the literature (Bastos et al. 2017) for the proportion who are dead at the End Of Treatment (EOT).

Using these estimates:

- Let $p(dead)_{o,EOT,MDR}$ be the probability that an MDR-TB patient (here, exclusive of XDR) will be observed as dead at the EOT. $p(dead)_{o,EOT,MDR} = 0.08,\ 95\%\ CI : (0.07, 0.09)$.
- Let $p(dead)_{o,EOT,XDR}$ be the probability that an XDR-TB patient will be observed as dead at the EOT. $p(dead)_{o,EOT,XDR} = 0.21,\ 95\%\ CI : (0.18, 0.25)$.

Based on the model mechanisms, we allow for truly cured individuals to have a lower mortality rate, which is a multiplier of background mortality. As a result, the overall mortality estimates from the literature are an **average**: at any given time point, the observed mortality rate is assumed to be a mean mortality rate of a two groups, one (lower) mortality rate in the cured group, and another (higher) mortality rate in the non-cured group. The challenge is that the proportion cured varies over time. The following details the approach we have taken, which is applied to the MDR-only and XDR population separately.

Let $\mu_{o,T} = -ln(1 - p(dead)_{o,EOT})$ be the observed mortality rate in the population over the whole treatment course, converted from the observed proportion dead at the EOT.

Let $\mu_C$ be the monthly mortality rate among those truly cured. This does not vary by month.

Let $\mu_{NC}$ be the monthly mortality rate among those not truly cured. This does not vary by month.

Let $t \in \{1, 2, \ldots, 21\}$ denote the month of the regimen. We assume for these calculations that EOT outcomes are recorded at 21 months, the modal regimen duration in the studies comprising the Bastos et al. 2017 review.

Let $p(cured)_t$ be the proportion of alive patients truly cured at the end of month $t$.

Among alive patients, true cure and true non-cure are mutually exclusive and collectively exhaustive. As such, $p(cured)_t + p(\overline{cured})_t = 1,\ \forall t$.

Then,

$$\mu_{o,T} = \sum_{t=1}^{T} [\mu_C * p(cured)_t + \mu_{NC} * p(\overline{cured})_t]$$

$$\mu_{o,T} = \sum_{t=1}^{T} [\mu_C * p(cured)_t + \mu_{NC} * (1 - p(cured)_t)]$$

$$\mu_{o,T} = \sum_{t=1}^{T} [\mu_C * p(cured)_t + \mu_{NC} - \mu_{NC} * p(cured)_t)]$$

$$\mu_{o,T} = T\mu_{NC} + \sum_{t=1}^{T} [\mu_C * p(cured)_t - \mu_{NC} * p(cured)_t)]$$

$$\mu_{o,T} = T\mu_{NC} + \sum_{t=1}^{T} [\mu_C * p(cured)_t] - \sum_{t=1}^{T} [\mu_{NC} * p(cured)_t)]$$

$$\mu_{o,T} - \sum_{t=1}^{T} [\mu_C * p(cured)_t] = T\mu_{NC} - \sum_{t=1}^{T} [\mu_{NC} * p(cured)_t)]$$

$$\mu_{o,T} - \mu_C \sum_{t=1}^{T} p(cured)_t = T\mu_{NC} - \mu_{NC} \sum_{t=1}^{T} p(cured)_t)$$

$$\mu_{o,T} - \mu_C \sum_{t=1}^{T} p(cured)_t = \mu_{NC}(T - \sum_{t=1}^{T} p(cured)_t))$$

$$\mu_{NC} = \frac{\mu_{o,T} - \mu_C \sum_{t=1}^{T} p(cured)_t}{(T - \sum_{t=1}^{T} p(cured)_t))} \qquad (1)$$

We are left with a formula with two unknowns, $\mu_{NC}$ and $p(cured)_t$.

The next step is to provide an approximation for $p(cured)_t$, the proportion of alive patients who are cured at time $t$.

Let $p(cured)_0 = 0$.

Let $p(cured)_1 = 0.019$, allowing only for the mechanism of self-cure in month 1.

Let $p(cured)_{21}$ be the proportion of patients cured among those alive from the Bastos et al 2017 review. As such, for MDR-TB patients, this will be approximated by a numerator of the observed proportion of treatment successes at end of treatment, and a denominator of the proportion alive at the end of treatment:

$$p(cured)_{MDR,21} = \frac{0.64}{1-0.08} = 0.70$$

For XDR-TB patients, this is approximated in the same way:

$$p(cured)_{XDR,21} = \frac{0.26}{1-0.21} = 0.33$$

To obtain the proportion cured among those alive for months 2-20 in these calculations, we make the simplifying assumption of a constant rate - among the alive non-cured - of converting to cure. We also ignore the dual mechanisms of self-cure and treatment-related cure, along with any differences in duration between MDR- and XDR-TB regimens:

As such, we obtain a monthly cure rate for alive MDR-TB patients, $\mu_{cure,MDR}$ as follows:

$$\mu_{cure,MDR} = \frac{-ln(1-p(cured)_{MDR,21})}{21}$$

$$\mu_{cure,MDR} = \frac{-ln(1-0.70)}{21}$$

$$\mu_{cure,MDR} = 0.057$$

Similarly, we obtain the monthly cure rate for alive XDR-TB patients, $\mu_{cure,XDR}$ as follows,

$$\mu_{cure,XDR} = \frac{-ln(1-p(cured)_{XDR,21})}{21}$$

$$\mu_{cure,XDR} = \frac{-ln(1-0.33)}{21}$$

$$\mu_{cure,XDR} = 0.019$$

From these, we can obtain the cumulative proportion of alive patients cured during each month (not shown). We plug these into formula (1),

$$\mu_{NC} = \frac{\mu_{o,T} - \mu_C \sum_{t=1}^{T} p(cured)_t}{(T - \sum_{t=1}^{T} p(cured)_t)}$$

$T = 21$

For MDR-TB,

$\mu_{o,T,MDR} = -ln(1 - 0.08)$

$\mu_{o,T,MDR} = 0.083$

For XDR-TB,

$\mu_{o,T,XDR} = -ln(1 - 0.21)$

$\mu_{o,T,XDR} = 0.236$

$\mu_C$, the monthly mortality rate among the cured, is a function of background (ASR) mortality. The median age was 35 in the Bastos et al. 2017 review, and the modal country of included studies was South Africa, which is also approximately middle of the pack for the income level of the meta-analysis. As such, we adopt the monthly mortality rate for a 35 year old individual in South Africa, and the mortality rate ratio ($MRR$) for MDR-TB from the parameter table.

$\mu_{ASR} = 0.00069$

$MRR = 3.07$

$\mu_C = \mu_{ASR} * MRR$

$\mu_C = 0.00069 * 3.07$

$\mu_C = 0.00211$

The mortality rate among those cured is assumed for now to be the same among those with MDR-TB and XDR-TB, although this could be edited going forward.

From the Excel table "Estimating mort rates", we have the following values too:

$\sum_{t=1}^{T} p(cured)_{t,MDR} = 9.102$

$\sum_{t=1}^{T} p(cured)_{t,MDR} = 3.519$

Putting all of this together, for MDR-TB:

$$\mu_{NC,MDR} = \frac{\mu_{o,T,MDR} - \mu_C \sum_{t=1}^{T} p(cured)_{t,MDR}}{(T - \sum_{t=1}^{T} p(cured)_{t,MDR}))}$$

$$\mu_{NC,MDR} = \frac{0.083 - 0.00211 * 9.102}{(21 - 9.102))}$$

$$\mu_{NC,MDR} = 0.00536$$

And for XDR-TB,

$$\mu_{NC,XDR} = \frac{\mu_{o,T,XDR} - \mu_C \sum_{t=1}^{T} p(cured)_{t,XDR}}{(T - \sum_{t=1}^{T} p(cured)_{t,XDR}))}$$

$$\mu_{NC,XDR} = \frac{0.236 - 0.00211 * 3.519}{(21 - 3.519))}$$

$$\mu_{NC,XDR} = 0.01307$$
