## Supplemental Figure 9 for "Impact and cost-effectiveness of the 6-month BPaLM regimen for rifampicin-resistant tuberculosis: a mathematical modeling analysis"

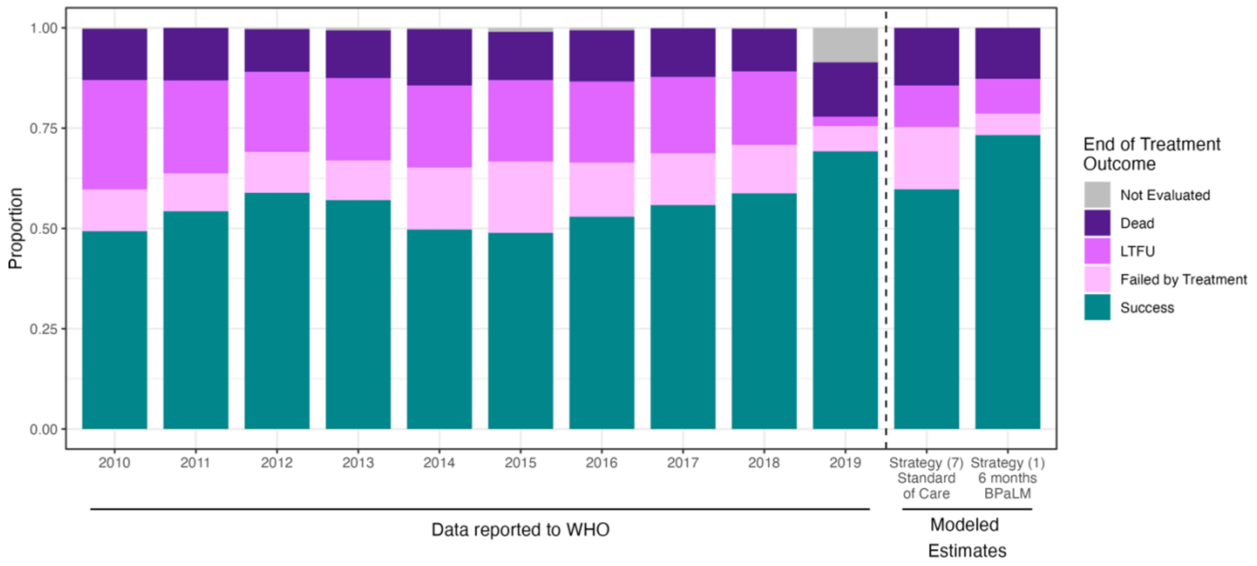


**S9 Fig. Validating modeled End of Treatment outcomes against data reported to WHO.**

The proportions recorded for each EOT outcome are shown for WHO RR-TB data 2010–2019 for Moldova (left of the vertical dashed line) and the modeled cohort outcomes (right of the vertical dashed line), where we assume that all EOT outcome categories are mutually exclusive, and that death during treatment or LTFU take precedence over a preceding treatment failure. Standard of care refers to modelled Strategy 7, and 6 months BPaLM refers to modelled Strategy 1. The number of observations per year in the WHO TB outcomes data for all MDR/RR-TB is in the range (559, 996).

BPaLM – bedaquiline, pretomanid, linezolid, moxifloxacin; EOT – End Of Treatment; LTFU – Lost to Follow Up; RR-TB – Rifampicin-resistant tuberculosis; WHO – World Health Organization moxifloxacin; FQ-R – fluoroquinolone-resistant; FQ-S – fluoroquinolone-susceptible; Mfx – moxifloxacin; QALY – quality-adjusted life year; USD – United States dollars; WTP – willingness-to-pay
