## Supplemental Figure 7 for "Impact and cost-effectiveness of the 6-month BPaLM regimen for rifampicin-resistant tuberculosis: a mathematical modeling analysis"

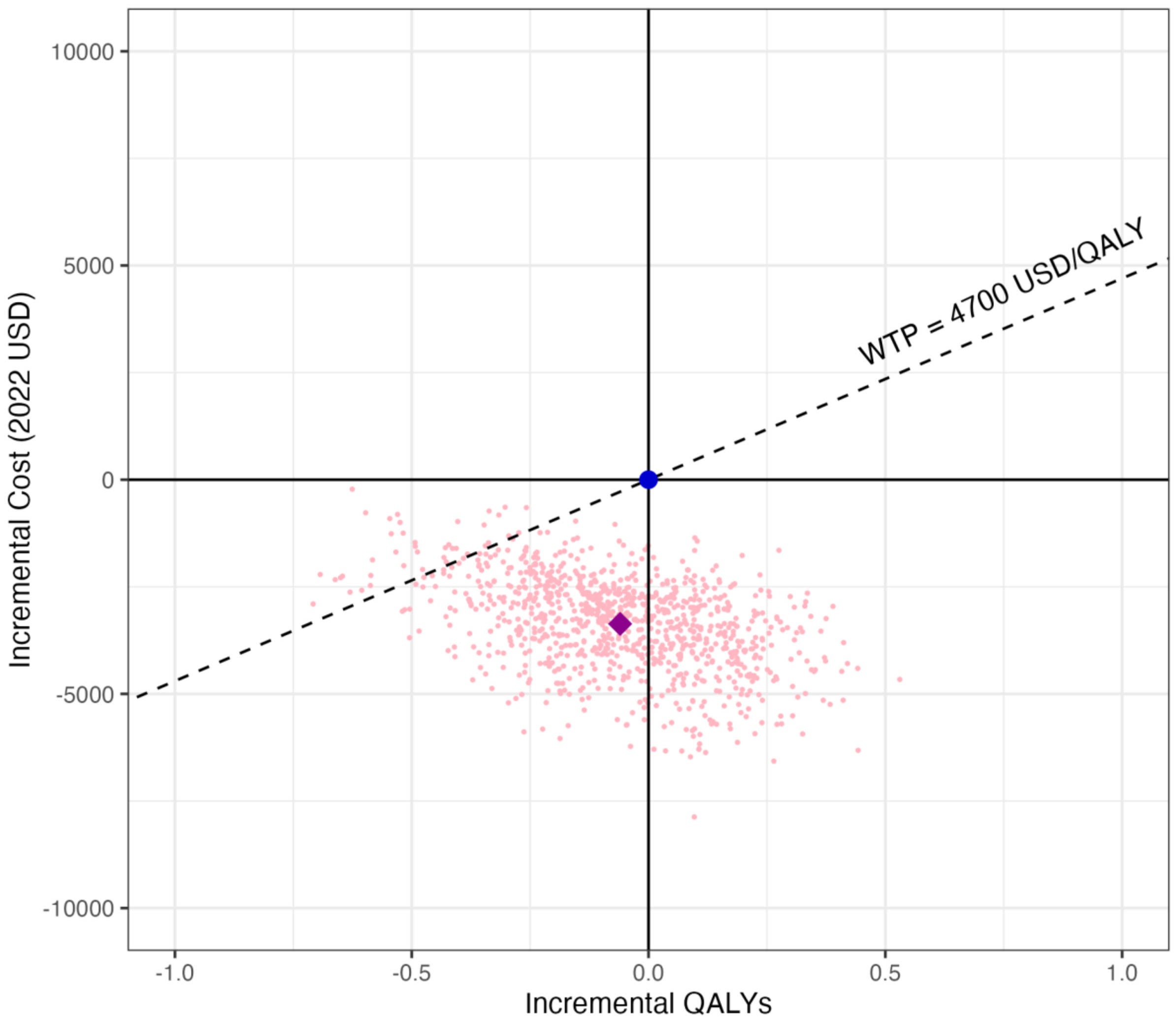


**S7 Fig. Incremental cost-effectiveness plane for the leading 6-month BPaLM strategy vs. the leading SOC strategy.**

The incremental cost-effectiveness plane compares the incremental discounted total QALYs and incremental discounted total costs for Strategy (1) as compared to a reference of Strategy (7). Each light pink point represents one iteration of the second-order Monte Carlo simulation, itself an average of 10,000 individual patient simulations. The purple diamond is the mean of the 1,000 second-order Monte Carlo simulations, corresponding to the point estimates in Table 3. The blue dot represents the standard of care (Strategy (7)), which is the reference point. BPaL – bedaquiline, pretomanid, linezolid; BPaLC – bedaquiline, pretomanid, linezolid, clofazimine; BPaLM – bedaquiline, pretomanid, linezolid, moxifloxacin; FQ-R – fluoroquinolone-resistant; FQ-S – fluoroquinolone-susceptible; Mfx – moxifloxacin; QALY – quality-adjusted life year; USD – United States dollars; WTP – willingness-to-pay
