## Supplemental Figure 6 for "Impact and cost-effectiveness of the 6-month BPaLM regimen for rifampicin-resistant tuberculosis: a mathematical modeling analysis"

A

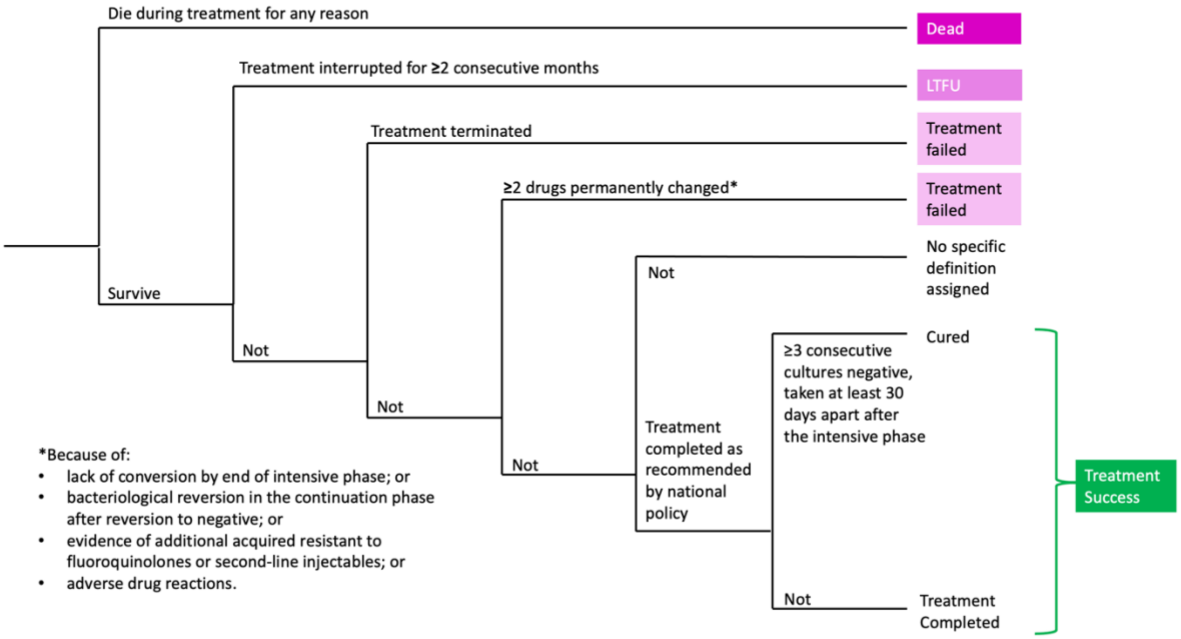


B

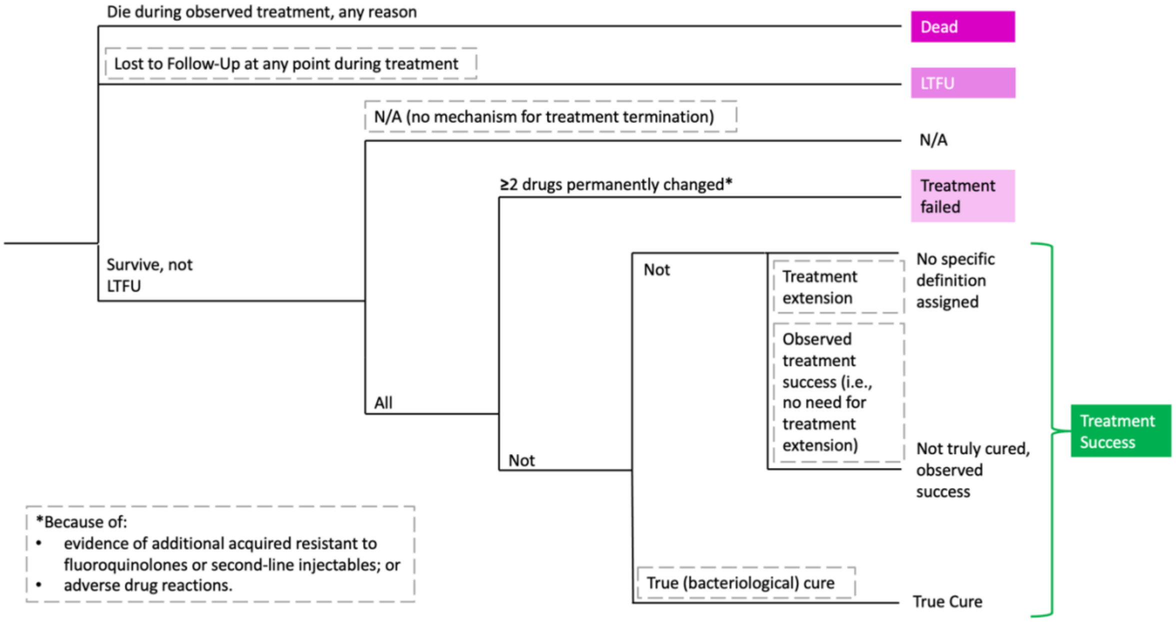


**S6 Fig. WHO-based definitions for End of Treatment Outcomes.**

Schematic showing the definitions for end of treatment outcomes used by the WHO (A), and this model (B). The constituents of each of the major end of treatment outcome categories are shown, as applied to all RR-TB including MDR-TB and XDR-TB. Differences between the WHO definitions and those used in this model are highlighted by the gray hashed boxes. The WHO definitions are not necessarily mutually exclusive; we assumed that the classification takes place according to the tree structure in (A), and implemented the aligned structure in (B) for tractability given the model mechanisms. For example, an individual who failed treatment and then died would be recorded as a death, because the branch involving death is closer to the root of the tree.
