## Supplemental Figure 4 for "Impact and cost-effectiveness of the 6-month BPaLM regimen for rifampicin-resistant tuberculosis: a mathematical modeling analysis"

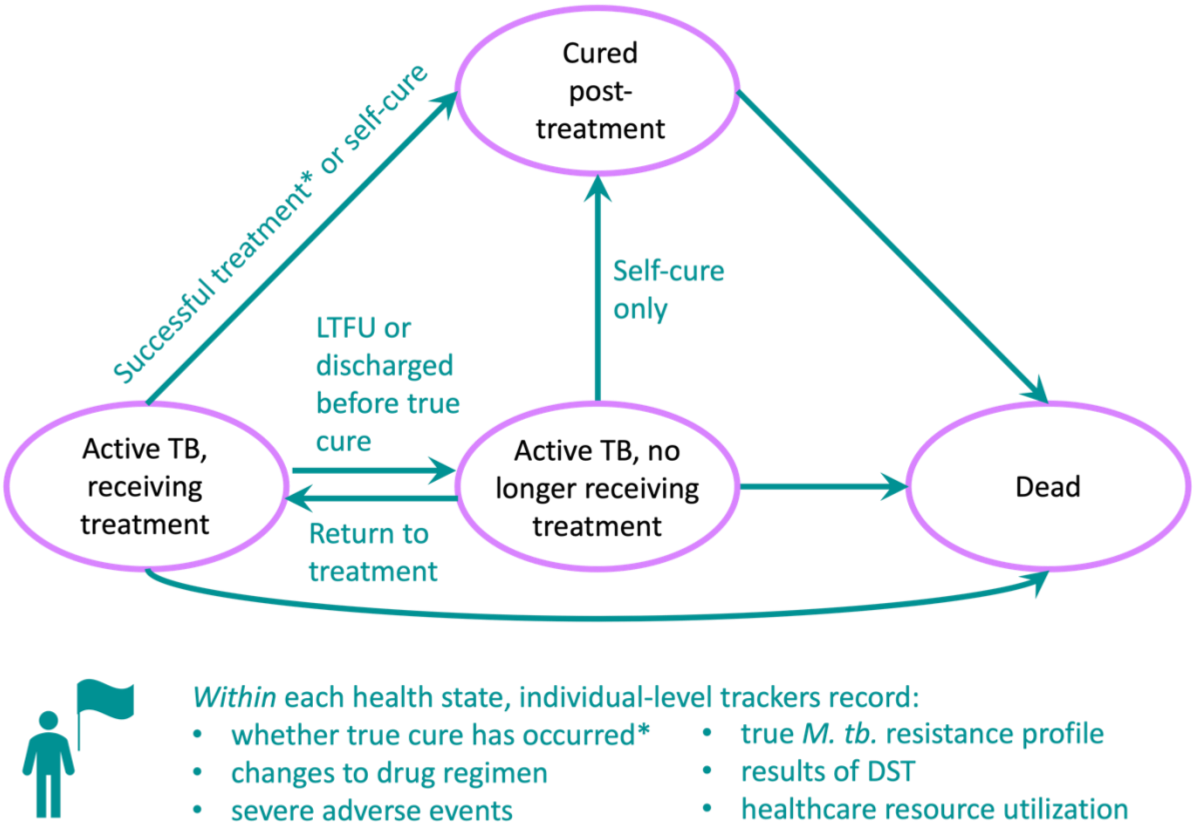


**S4 Fig. Markov state-transition diagram.**

Transitions between states can occur as shown by the arrows. Though not receiving treatment, individuals in the "Active TB, no longer receiving treatment" state are subject to a low rate of self-cure, and so may still transition to the "Cured post-treatment" state. Asterisks (*) highlight the major mechanisms through which the choice of treatment intervention affects outcomes. LTFU – Lost to follow-up; TB – tuberculosis.
