## Supplemental Figure 3 for "Impact and cost-effectiveness of the 6-month BPaLM regimen for rifampicin-resistant tuberculosis: a mathematical modeling analysis"

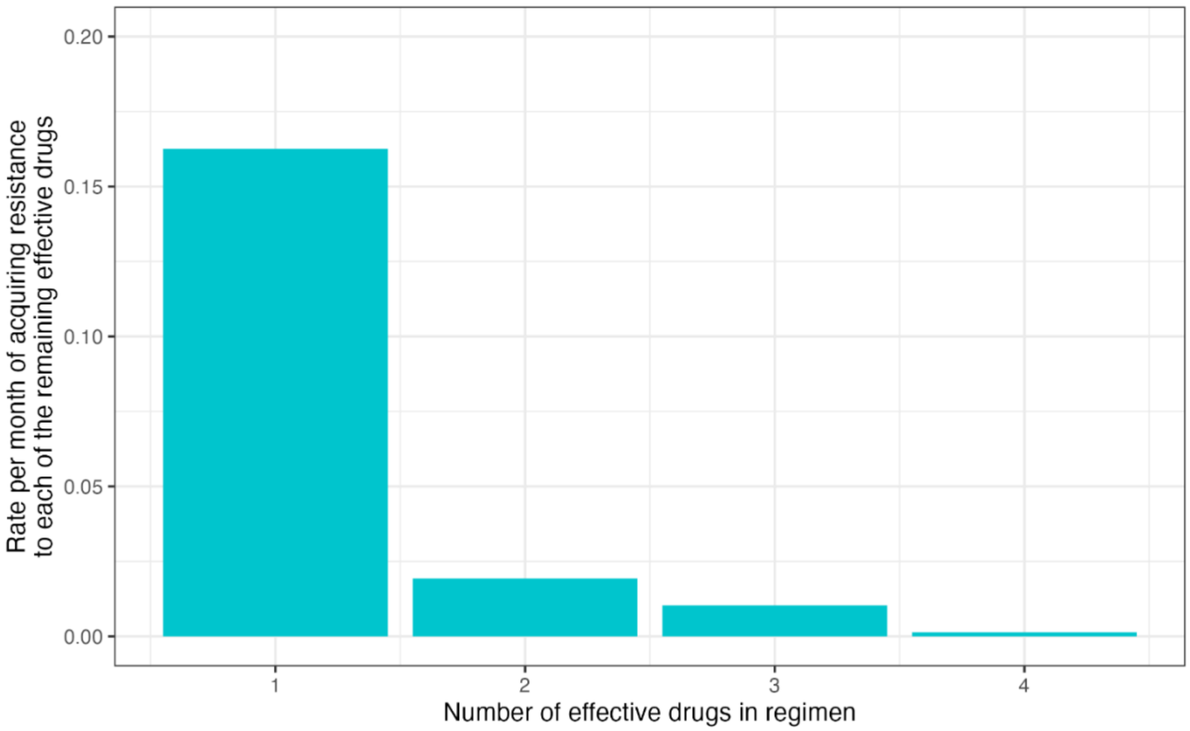


**S3 Fig. The rate of acquiring drug resistance.**

The modeled point estimate for the monthly rate that an individual's strain of *M. tuberculosis* will acquire resistance to each effective drug it is exposed to is plotted, conditional on that individual beginning the month with *n* effective drugs in the regimen (x-axis). Estimates for 1, 3 and 4 effective drugs were obtained from the literature. The estimate for 2 drugs was calculated, assuming an additive risk (i.e., the increase in risk for 2 effective drugs compared to 3 is the same as the increase in risk for 3 effective drugs compared to 4). See also S1 Table.
