## Supplemental Figure 8 for "Impact and cost-effectiveness of the 6-month BPaLM regimen for rifampicin-resistant tuberculosis: a mathematical modeling analysis"

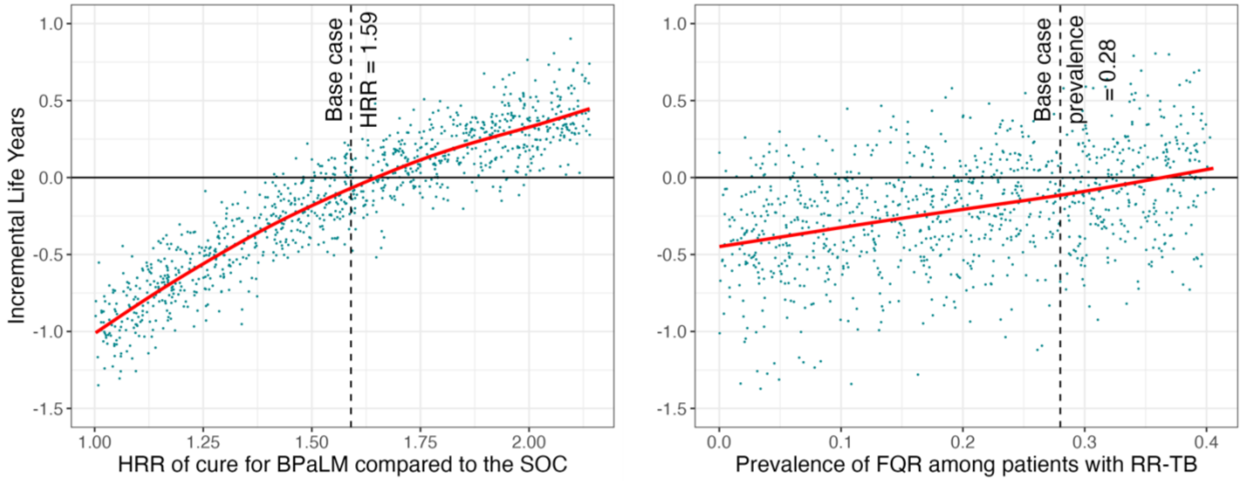


**S8 Fig. Sensitivity analyses varying relative effectiveness of BPaLM and cohort prevalence of fluoroquinolone resistance, outcome of Life Years.**

One-way sensitivity analyses testing the effect of key model parameter assumptions, in terms of their effect on the incremental Life Years experienced under the Strategy (1) (6 months BPaLM, DST upfront, repeat DST every 4 months, BPaLC if Mfx stopped) as compared to Strategy (7) (standard of care 9-18 month regimens based on results of upfront DST, repeat DST every 4 months). We chose to compare these two strategies as they were the best-performing BPaLM-based and standard of care-based strategies, respectively. Each of the parameters is varied deterministically in the respective sensitivity analysis, with all other model parameters drawn as in the probabilistic sensitivity analysis. In the left column, the HRR of cure for the BPaLM regimen compared to the standard of care is varied. In the right column, we vary the starting prevalence of fluoroquinolone resistance in the cohort. Each of 1,000 model runs is shown in each plot, itself an average of 10,000 individual patient simulations. The red line shows the trend as represented by regression of the y-axis variable on the x-axis variable, using a generalized additive model with cubic spline to obtain a restricted maximum likelihood within ggplot2 [1]. The vertical dashed lines mark the base case assumption for the mean of each of these model parameters. FQR – Fluoroquinolone Resistance; HRR – Hazard Rate Ratio; LY –Life Year; RR-TB – Rifampicin-resistant tuberculosis
