## Supplemental Figure 5 for "Impact and cost-effectiveness of the 6-month BPaLM regimen for rifampicin-resistant tuberculosis: a mathematical modeling analysis"

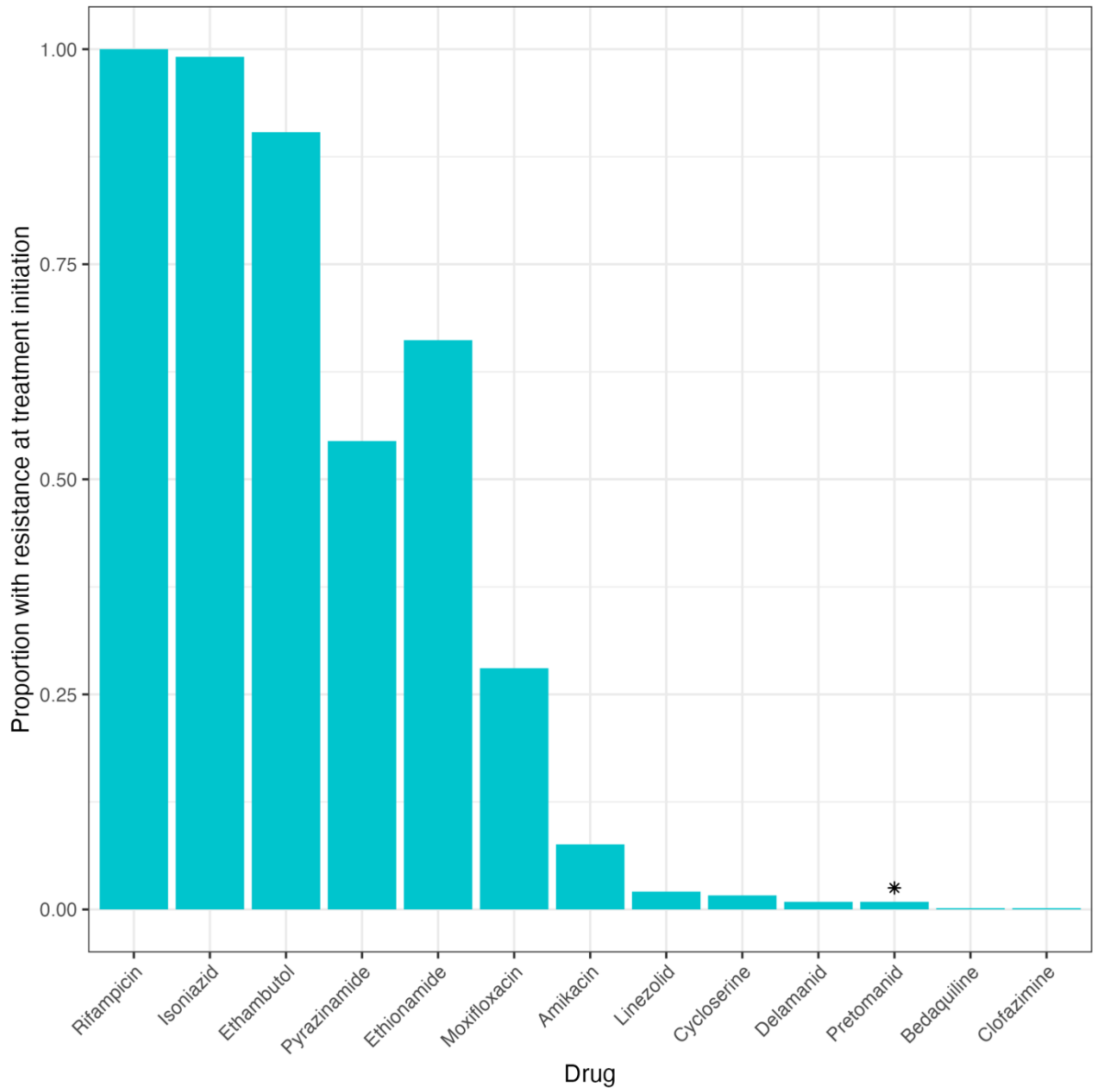


**S5 Fig. Cohort prevalence of *M. tb.* resistance to key drugs at treatment initiation.**

The proportion of the cohort with primary resistance to each drug is plotted, as described by *M. tuberculosis* whole genomic sequencing data from Moldova [1,2]. All those observations with rifampicin susceptibility were excluded, as per S3 Fig. *There was no resistance data for pretomanid; resistance was assumed to be at the same level as for delamanid.

**REFERENCES**

These references are provided here for convenience. They are also cited within the main manuscript file in the legend for S5 Fig.


1. ID 736718 - BioProject - NCBI. [date accessed: 9 Feb 2023]. Available: https://www-ncbi-nlm-nih-gov.ezp-prod1.hul.harvard.edu/bioproject/PRJNA736718

2. Yang C, Sobkowiak B, Naidu V, Codreanu A, Ciobanu N, Gunasekera KS, et al. Phylogeography and transmission of M. tuberculosis in Moldova. 2021 Jul p. 2021.06.30.21259748. doi:10.1101/2021.06.30.21259748
