## Supplemental Figure 2 for "Impact and cost-effectiveness of the 6-month BPaLM regimen for rifampicin-resistant tuberculosis: a mathematical modeling analysis"

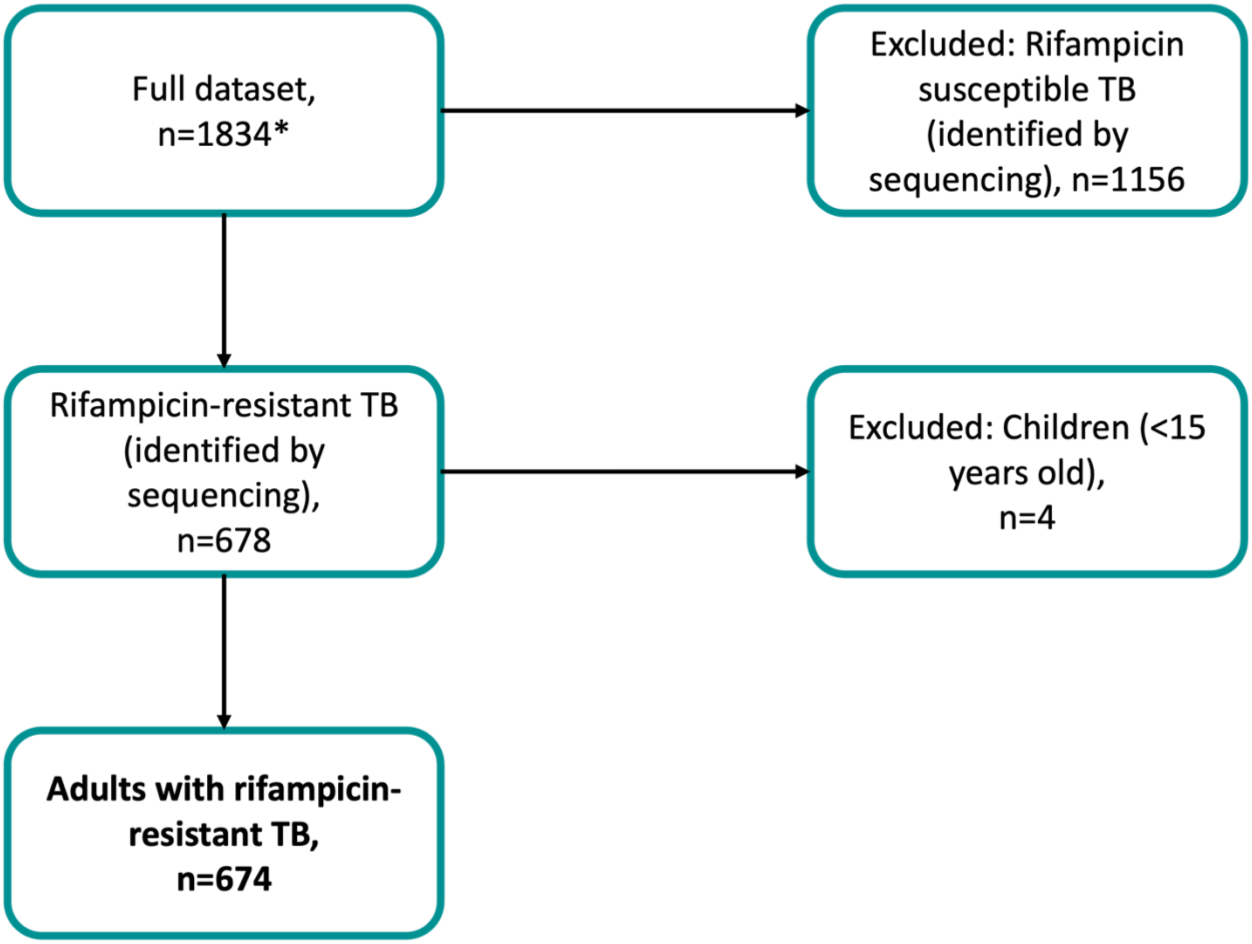


**S2 Fig**. ***M. tb.* genomic sequencing data exclusion criteria.**

*The full dataset did not include any samples with mixed strains of *M. tb.*

Exclusions made to the genomic sequencing drug susceptibility testing dataset are shown along with the number of observations. This dataset is described elsewhere [1,2]. The presence of a mutation conferring resistance to rifampicin was assumed to convey full resistance, and vice versa.

TB – tuberculosis

**REFERENCES**

These references are provided here for convenience. They are also cited within the main manuscript file in the legend for S2 Fig.

1. ID 736718 - BioProject - NCBI. [date accessed: 9 Feb 2023]. Available: https://www-ncbi-nlm-nih-gov.ezp-prod1.hul.harvard.edu/bioproject/PRJNA736718

2. Yang C, Sobkowiak B, Naidu V, Codreanu A, Ciobanu N, Gunasekera KS, et al. Phylogeography and transmission of M. tuberculosis in Moldova. 2021 Jul p. 2021.06.30.21259748. doi:10.1101/2021.06.30.21259748
