## Supplemental Figure 1 for "Impact and cost-effectiveness of the 6-month BPaLM regimen for rifampicin-resistant tuberculosis: a mathematical modeling analysis"

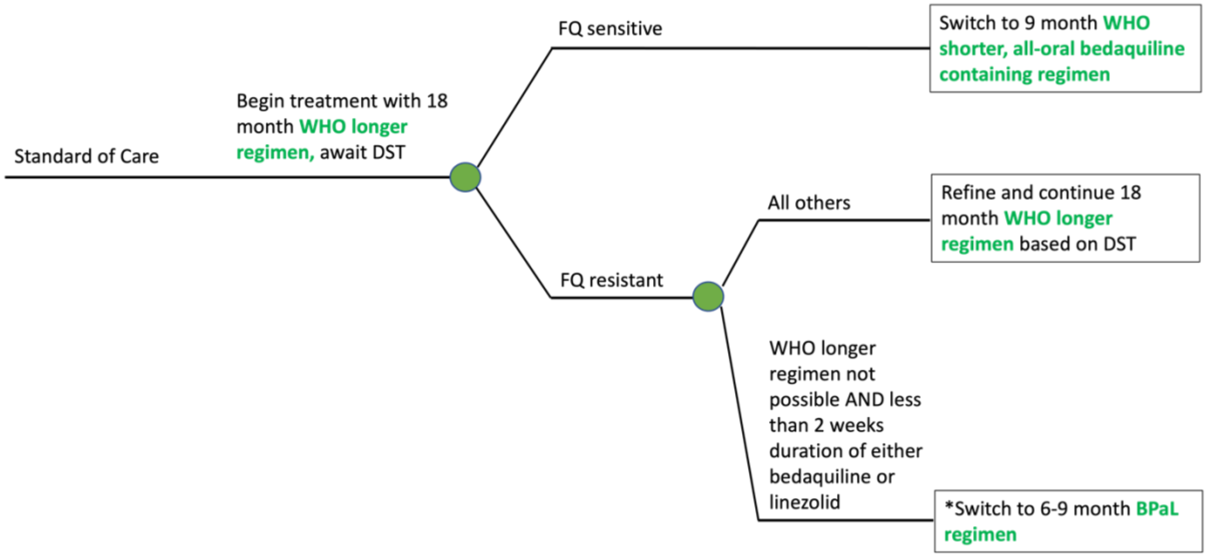


**S1 Fig. Schematic of the initial workup phase for the standard of care**

Both standard of care strategies (Strategy 7 and Strategy 8) are modeled on the recommended workup and regimen selection in the 2020 WHO guidelines on the treatment of drug-resistant tuberculosis [1]. We assumed that DST results (by MGIT) are available in 2 weeks. *While we include the BPaL regimen as per the guidelines, no patients actually met the criteria to receive it under the standard of care (Strategies 7 and 8) in our model (i.e., in all model simulations, it is possible to adopt a WHO longer regimen). BPaL – bedaquiline, pretomanid, linezolid; DST – drug susceptibility test; FQ – fluoroquinolone; MGIT – Mycobacterial Growth Indicator Tube; WHO – World Health Organization

**REFERENCE**

This references is provided here for convenience. It are also cited within the main manuscript file in the legend for S1 Fig.

1. WHO Consolidated Guidelines on Tuberculosis, Module 4: Treatment - Drug-Resistant Tuberculosis Treatment. World Health Organization; 2020. Available: https://www.who.int/publications-detail-redirect/9789240007048
