## Supplemental Table 6 for "Impact and cost-effectiveness of the 6-month BPaLM regimen for rifampicin-resistant tuberculosis: a mathematical modeling analysis"

**S6 Table. Five-year budget impact of implementing 6 months of BPaLM in Moldova.**

| **Budget Year** | **TB Program Budget Category** | | | | |
|---|---|---|---|---|---|
| | **Drugs** | **Laboratory Tests** | **Routine Care** | **Nonroutine Care** | **Total** |
| Year 1 | -583,795<br>(-1,275,027, -96,331)<br>p=0.002 | -842<br>(-2,946, 523)<br>p=0.282 | -47,267<br>(-106,039, -10,296)<br>p=0.002 | 380<br>(-738, 1,902)<br>p=0.588 | -631,524<br>(-1,352,836, -115,576)<br>p=0.002 |
| Year 2 | -1,271,079<br>(-2,780,357, -213,592)<br>p=0.002 | -2,206<br>(-8,253, 2,182)<br>p=0.36 | -122,422<br>(-288,184, -24,477)<br>p=0.002 | 391<br>(-1,719, 3,147)<br>p=0.786 | -1,395,316<br>(-3,008,507, -254,755)<br>p=0.002 |
| Year 3 | -1,475,708<br>(-3,254,882, -249,633)<br>p=0.002 | -3,328<br>(-12,732, 3,751)<br>p=0.396 | -159,279<br>(-395,607, -23,918)<br>p=0.006 | -1<br>(-2,287, 2,689)<br>p=0.93 | -1,638,316<br>(-3,532,616, -299,848)<br>p=0.002 |
| Year 4 | -1,529,185<br>(-3,367,264, -253,873)<br>p=0.002 | -4,283<br>(-15,682, 4,332)<br>p=0.364 | -172,308<br>(-445,490, -19,667)<br>p=0.014 | -186<br>(-2,543, 2,525)<br>p=0.802 | -1,705,962<br>(-3,694,619, -313,741)<br>p=0.002 |
| Year 5 | -1,555,715<br>(-3,421,880, -260,667)<br>p=0.002 | -5,044<br>(-17,929, 4,367)<br>p=0.326 | -183,809<br>(-485,833, -18,711)<br>p=0.024 | -306<br>(-2,693, 2,395)<br>p=0.722 | -1,744,873<br>(-3,794,268, -321,588)<br>p=0.002 |
| **Total** | -6,415,481<br>(-14,173,208, -1,077,963)<br>p=0.002 | -15,704<br>(-57,639, 14,895)<br>p=0.354 | -685,085<br>(-1,716,138, -97,308)<br>p=0.006 | 278<br>(-9,970, 12,677)<br>p=0.98 | -7,115,992<br>(-15,396,823, -1,301,637)<br>p=0.002 |

TB – tuberculosis.

The budget impact was estimated for 6 months BPaLM (Strategy (1)) as compared to standard of care (Strategy (7))
