## Supplemental Table 5 for "Impact and cost-effectiveness of the 6-month BPaLM regimen for rifampicin-resistant tuberculosis: a mathematical modeling analysis"

**S5 Table. Comparing outcomes at 6 months, 12 months, and 72 weeks from treatment initiation.**

| STRATEGY DETAILS | | | | RISK OF UNFAVORABLE OUTCOMES | | | |
|---|---|---|---|---|---|---|---|
| **Strategy Name** | **Alternative regimen if Mfx stopped** | **2nd-line DST at treatment initiation** | **Routine frequency of subsequent 2nd-line DST** | **Based on TB-PRACTECAL trial definition** | | **Based on WHO End of Treatment definition** | |
| | | | | Proportion (%) | Risk Difference (p.p.) | Proportion (%) | Risk Difference (p.p.) |
| *6 months from treatment initiation* | | | | | | | |
| (7) Standard of Care | -- | Yes | Every 4 months | 32.9 (30.1, 36) | -- | 29.6 (27, 32.6) | -- |
| (1) 6 months BPaLM | BPaLC | Yes | Every 4 months | 30.9 (27.7, 34.5) | -1.9 (-5.2, 1.4) p=0.234 | 17.8 (16, 20) | -11.8 (-14.6, -9.4) p=<0.001 |
| *12 months from treatment initiation* | | | | | | | |
| (7) Standard of Care | -- | Yes | Every 4 months | 39.0 (36.3, 42.2) | | 35.8 (33.1, 38.9) | |
| (1) 6 months BPaLM | BPaLC | Yes | Every 4 months | 36.1 (32.6, 39.8) | -2.9 (-5.8, 0.2) p=0.068 | 22.8 (20.4, 25.4) | -12.9 (-15.6, -10.5) p=<0.001 |
| *17 months (i.e., 72 weeks) from treatment initiation* | | | | | | | |
| (7) Standard of Care | -- | Yes | Every 4 months | 41.8 (39.1, 45.1) | | 38.0 (35.1, 41.3) | |
| (1) 6 months BPaLM | BPaLC | Yes | Every 4 months | 38.7 (35.2, 42.5) | -3.1 (-6.0, 0.0) p=0.050 | 24.5 (21.9, 27.5) | -13.4 (-16.1, -11.0) p=<0.001 |

BPaLC – bedaquiline, pretomanid, linezolid, clofazimine; BPaLM – bedaquiline, pretomanid, linezolid, moxifloxacin; LYs – Life Years; Mfx – moxifloxacin; p.p. – percentage points; QALYs – Quality-adjusted Life Years; UI – Uncertainty Interval; WHO – World Health Organization

The following health outcomes are shown: a composite “Unfavorable outcome” closely aligned to the composite trial endpoint in TB-PRACTECAL, true cure, and quality-adjusted life expectancy. Results are shown separately over three model-run time horizons: 6 months, 72 weeks (in line with the endpoint in TB-PRACTECAL), and lifetime (in line with the primary outcomes in our analysis).
