## Supplemental Table 4 for "Impact and cost-effectiveness of the 6-month BPaLM regimen for rifampicin-resistant tuberculosis: a mathematical modeling analysis"

**S4 Table. Life Years achieved under each RR-TB treatment strategy.**

| Strategy | Under BPaLM, alternative regimen if Moxifloxacin stopped | DST for second-line drugs at treatment initiation | Frequency of DST during subsequent treatment course | Undiscounted Life Years |
|---|---|---|---|---|
| 5) 6 months BPaLM | BPaLC | No | Every 4 months | 14.745 (12.72, 16.55) |
| 1) 6 months BPaLM | BPaLC | Yes | Every 4 months | 14.750 (12.76, 16.54) |
| 2) 6 months BPaLM | BPaLC | Yes | Monthly | 14.753 (12.74, 16.49) |
| 6) 6 months BPaLM | BPaL only | No | Every 4 months | 14.405 (12.44, 16.10) |
| 3) 6 months BPaLM | BPaL only | Yes | Every 4 months | 14.414 (12.42, 16.17) |
| 4) 6 months BPaLM | BPaL only | Yes | Monthly | 14.408 (12.42, 16.16) |
| 7) standard of care | -- | Yes | Every 4 months | 14.832 (13.00, 16.54) |
| 8) standard of care | -- | Yes | Monthly | 14.836 (12.98, 16.56) |

BPaL – bedaquiline, pretomanid, linezolid; BPaLC – bedaquiline, pretomanid, linezolid, clofazimine; BPaLM – bedaquiline, pretomanid, linezolid, moxifloxacin; UI – Uncertainty Interval.

Strategies are listed in the same order as Table 3. Mean values are shown with accompanying 95% UIs in parentheses.
