## Supplemental Table 3 for "Impact and cost-effectiveness of the 6-month BPaLM regimen for rifampicin-resistant tuberculosis: a mathematical modeling analysis"

### S3 Table. Duration and cumulative incidence of resistance to key drugs.

| Drugs | Duration with resistance, 6 months BPaLM, entire cohort (months) | Duration with resistance, SOC, entire cohort (months) | Difference in duration, BPaLM vs. SOC, entire cohort | p-value | Duration with resistance, 6 months BPaLM, active untreated TB (months) | Duration with resistance, SOC, active untreated TB (months) | Difference in duration, BPaLM vs. SOC, active untreated TB | p-value | Lifetime cumulative incidence of resistance, 6 months BPaLM (%) | Lifetime cumulative incidence of resistance, SOC (%) | Difference in cumulative incidence, BPaLM vs. SOC (p.p.) | p-value |
|---|---|---|---|---|---|---|---|---|---|---|---|---|
| Amikacin | 1.79 (1.4, 2.25) | 2.18 (1.66, 2.71) | -0.4 (-0.79, -0.06) | 0.022 | 0.43 (0.31, 0.56) | 0.42 (0.29, 0.55) | 0.01 (-0.1, 0.12) | 0.868 | 0 (0, 0) | 0 (0, 0.02) | 0 (-0.02, 0) | <0.001 |
| Bedaquiline | 0.93 (0.56, 1.47) | 1.86 (1.29, 2.51) | -0.92 (-1.48, -0.49) | <0.001 | 0.21 (0.12, 0.34) | 0.39 (0.26, 0.54) | -0.18 (-0.3, -0.06) | <0.001 | 3.13 (2.22, 4.33) | 5.22 (4.07, 6.59) | -2.09 (-3.03, -1.24) | <0.001 |
| Clofazimine | 1.09 (0.66, 1.67) | 2.4 (1.75, 3.2) | -1.31 (-1.94, -0.8) | <0.001 | 0.24 (0.15, 0.39) | 0.5 (0.35, 0.68) | -0.25 (-0.4, -0.12) | <0.001 | 3.81 (2.71, 5.17) | 7.67 (6.1, 9.55) | -3.86 (-5.05, -2.8) | <0.001 |
| Cycloserine | 0.61 (0.42, 0.88) | 1.56 (1.15, 2.05) | -0.95 (-1.38, -0.62) | <0.001 | 0.14 (0.09, 0.21) | 0.31 (0.22, 0.44) | -0.17 (-0.28, -0.09) | <0.001 | 0.75 (0.45, 1.15) | 3.07 (2.29, 3.97) | -2.32 (-3.1, -1.66) | <0.001 |
| Delamanid | 1.06 (0.65, 1.6) | 0.52 (0.37, 0.7) | 0.54 (0.18, 1.04) | 0.002 | 0.24 (0.14, 0.37) | 0.11 (0.07, 0.16) | 0.13 (0.04, 0.24) | <0.001 | 3.16 (2.2, 4.32) | 1.5 (0.98, 2.13) | 1.66 (0.92, 2.52) | <0.001 |
| Ethambutol | 13.48 (10.92, 16.5) | 15.01 (13.75, 16.16) | -1.52 (-4.26, 1.45) | 0.302 | 3.26 (2.58, 4.13) | 2.99 (2.6, 3.41) | 0.27 (-0.4, 1.02) | 0.474 | 0 (0, 0) | 0.19 (0.1, 0.3) | -0.19 (-0.3, -0.1) | <0.001 |
| Ethionamide | 9.92 (8.05, 12.08) | 11.39 (10.42, 12.29) | -1.47 (-3.47, 0.7) | 0.202 | 2.4 (1.88, 3.05) | 2.28 (1.96, 2.59) | 0.12 (-0.38, 0.67) | 0.714 | 0 (0, 0) | 0.37 (0.2, 0.56) | -0.37 (-0.56, -0.2) | <0.001 |
| Isoniazid | 14.76 (11.92, 18.09) | 15.91 (14.57, 17.11) | -1.16 (-4.11, 2.18) | 0.466 | 3.57 (2.81, 4.53) | 3.16 (2.73, 3.59) | 0.41 (-0.32, 1.23) | 0.294 | 0 (0, 0) | 0.01 (0, 0.03) | -0.01 (-0.03, 0) | <0.001 |
| Linezolid | 0.93 (0.64, 1.29) | 1.15 (0.85, 1.53) | -0.22 (-0.54, 0.08) | 0.152 | 0.21 (0.14, 0.31) | 0.22 (0.15, 0.31) | -0.01 (-0.09, 0.07) | 0.77 | 1.25 (0.76, 1.87) | 1.15 (0.78, 1.57) | 0.1 (-0.33, 0.55) | 0.694 |
| Moxifloxacin | 5.41 (4.38, 6.73) | 7.62 (6.66, 8.65) | -2.21 (-3.39, -1.02) | <0.001 | 1.29 (0.99, 1.65) | 1.39 (1.15, 1.65) | -0.1 (-0.4, 0.22) | 0.548 | 1.72 (1.08, 2.5) | 1.47 (0.94, 2.09) | 0.25 (-0.19, 0.8) | 0.354 |
| Pretomanid | 1.06 (0.65, 1.6) | 0.51 (0.36, 0.69) | 0.55 (0.2, 1.05) | <0.001 | 0.24 (0.14, 0.37) | 0.11 (0.07, 0.16) | 0.13 (0.04, 0.24) | <0.001 | 3.16 (2.2, 4.32) | 1.4 (0.92, 1.98) | 1.76 (1.01, 2.66) | <0.001 |
| Pyrazinamide | 8.54 (6.92, 10.41) | 10.83 (9.84, 11.84) | -2.28 (-4.02, -0.52) | 0.016 | 2.06 (1.6, 2.59) | 2.12 (1.82, 2.44) | -0.06 (-0.51, 0.4) | 0.758 | 0 (0, 0) | 0.66 (0.4, 0.97) | -0.66 (-0.97, -0.4) | <0.001 |
| Rifampicin | 14.88 (12.03, 18.23) | 15.98 (14.63, 17.19) | -1.1 (-4.07, 2.28) | 0.486 | 3.6 (2.84, 4.57) | 3.17 (2.74, 3.61) | 0.43 (-0.31, 1.27) | 0.28 | N/A | N/A | N/A | N/A |

BPaLM, bedaquiline, pretomanid, linezolid, moxifloxacin; SOC, standard of care.

The entire cohort had RR-TB, and so the duration with rifampicin resistance is equivalent to the duration with active RR-TB, and the cumulative incidence of rifampicin resistance is not applicable. Some drugs were used very sparingly, if ever, under one or both strategies (e.g., amikacin, ethambutol, ethionamide, isoniazid, and pyrazinamide); as such the cumulative incidence may be very low for these drugs under one or both strategies.
