## Supplemental Table 2 for "Impact and cost-effectiveness of the 6-month BPaLM regimen for rifampicin-resistant tuberculosis: a mathematical modeling analysis"

**S2 Table. Probability of loss to follow up by month of treatment.**

| Time, t, in months | Proportion LTFU at *t* | LTFU-free survival at *t* | p(LTFU by *t+1* \| alive at *t*) | Adjusted* p(LTFU by *t+1* \| alive at *t*) |
|---|---|---|---|---|
| 0 | 0 | 1 | 0.025 | 0.025 |
| 1 | 0.0254 | 0.97461 | 0.009 | 0.009 |
| 2 | 0.0340 | 0.96601 | 0.012 | 0.012 |
| 3 | 0.0452 | 0.95481 | 0.011 | 0.011 |
| 4 | 0.0553 | 0.94472 | 0.015 | 0.015 |
| 5 | 0.0693 | 0.93072 | 0.015 | 0.015 |
| 6 | 0.0836 | 0.91641 | 0.013 | 0.013 |
| 7 | 0.0951 | 0.90487 | 0.015 | 0.015 |
| 8 | 0.1088 | 0.8912 | 0.011 | 0.011 |
| 9 | 0.1189 | 0.88114 | 0.008 | 0.008 |
| 10 | 0.1259 | 0.87414 | 0.005 | 0.005 |
| 11 | 0.1302 | 0.86977 | 0.011 | 0.011 |
| 12 | 0.1400 | 0.85997 | 0.005 | 0.005 |
| 13 | 0.1443 | 0.85567 | 0.005 | 0.005 |
| 14 | 0.1485 | 0.85146 | 0.008 | 0.008 |
| 15 | 0.1556 | 0.84436 | 0.003 | 0.003 |
| 16 | 0.1586 | 0.84145 | 0.005 | 0.005 |
| 17 | 0.1627 | 0.83731 | 0.002 | 0.002 |
| 18 | 0.1642 | 0.83577 | 0.005 | 0.005 |
| 19 | 0.1685 | 0.83154 | 0.003 | 0.003 |
| 20 | 0.1712 | 0.82885 | 0.000 | 0.000 |
| 21 | 0.1712 | 0.82885 | 0.000 | 0 |
| 22 | 0.1712 | 0.82882 | 0.001 | 0 |
| 23 | 0.1724 | 0.82765 | 0.000 | 0 |
| 24 | 0.1720 | 0.828 | -- | 0 |

LTFU – Lost to Follow Up.

LTFU data from Walker et al. 2019 [1]. *The values in the rightmost column are used as the model inputs. Compared to the fourth column, we rounded down the values from month 21 onwards such that the probability of LTFU is zero thenceforth.
