## Supplemental Table 1 for "Impact and cost-effectiveness of the 6-month BPaLM regimen for rifampicin-resistant tuberculosis: a mathematical modeling analysis"

**S1 Table. Model input parameters, complete set.**

| # | Parameter name | Point Estimate | Distribution | Source(s) | Notes |
|---|---|---|---|---|---|
| **TB NATURAL HISTORY** | | | | | |
| 1 | Probability of death from background causes, per year | Varies; see Notes column | N/A | UN Population Division 2019 [1] File MORT/15-1 Both Sexes. | Estimates for 2015-2020 period. Converted to monthly rates, assuming a constant mortality rate during each 1-year period. The 1 year probability of death is as follows: Age 18-20: 0.000413; age 20-25: 0.000559; age 25-30: 0.000841; age 30-35: 0.001294; age 35-40: 0.002320; age 40-45: 0.003268; age 45-50: 0.005827; age 50-55: 0.008702; age 55-60: 0.013162; age 60-65: 0.022045; age 65-70: 0.029006; age 70-75: 0.046571; age 75-80: 0.073896; age 80-85: 0.116146; age 85-90: 0.175700; age 90-95: 0.242534; age 95+: 0.330946 |
| 2 | Rate of death from untreated TB, annual | 0.389 | Published point estimate (median) and 95% CrI (0.335-0.449) modeled as Lognormal (mu -0.9442, sigma 0.0763) | Ragonnet R, et al. Clin Infect Dis 2020 [2] | Applied to those with TB who are no longer receiving treatment (i.e., those LTFU and those who appeared to successfully complete treatment but had not been truly cured). This estimate is for smear positive individuals; we discuss this as a limitation in the main text. |
| 2b | Rate of death from untreated TB, among those discharged prior to true cure, annual | 0.025 | Published point estimate (median) and 95% CrI (0.017, 0.035) modeled as Lognormal (mu -3.689, sigma 0.179) | Ragonnet R, et al. Clin Infect Dis 2020 [2] | Applied for first 98 months (see parameter #14 in this table). |
| 3 | Mortality rate ratio for those who are cured, compared to background mortality | 3.070 | Published point estimate and 95% CI (2.12, 4.45) modeled as Lognormal (mu 1.122, sigma 0.1889) | Romanowski K., et al. Lancet Infect Dis 2019 [3] | Estimate for pulmonary TB. Applied as a mortality rate ratio, although it was reported as a standardized mortality ratio (some of the constituent studies in the review had used mortality rates and some had used SMR). |

| # | Parameter name | Point Estimate | Distribution | Source(s) | Notes |
|---|---|---|---|---|---|
| 4 | Rate of self-cure, annual | 0.231 | Published point estimate and 95% CrI (0.177, 0.288) modeled as Lognormal (mu - 1.465, sigma 0.136) | Ragonnet R, et al. Clin Infect Dis 2020 [2] | Applied to those not receiving treatment (i.e., those LTFU and those who appeared to successfully complete treatment but had not been truly cured) and the first two months of treatment. |
| **TREATMENT-RELATED PARAMETERS** | | | | | |
| 5 | Probability of all-cause death for WHO longer regimen, MDR only (excluding XDR), at 21 months | 0.080 | N/A | Bastos M. L., et al. 2017 [4] | Estimate from a large meta-analysis. The regimen duration was variable across studies, typically in the range 18-24 months. To obtain a monthly estimate for disease-specific mortality (parameter [7]), we assumed a 21 month regimen duration. Further detail in Supplementary Material E: "Calculated Model Parameters". |
| 6 | Probability of all-cause death for WHO longer regimen, XDR only, at 21 months | 0.21 | N/A | Bastos M. L., et al. 2017 Table S3 [4] | Estimate from a large meta-analysis. The regimen duration was variable across studies, typically in the range 18-24 months. To obtain a monthly estimate for disease-specific mortality (parameter [8]), we assumed a 21 month regimen duration. Further detail in Supplementary Material E: "Calculated Model Parameters". |
| 7 | Mortality rate among those who are not cured but on treatment, MDR-TB only (excluding XDR-TB), monthly | 0.00536 | Beta (mean 0.00536, s.d. 0.00178)* | Bastos M. L., et al. 2017 [4] | See Supplementary Material E "Calculated Model Parameters". |

| # | Parameter name | Point Estimate | Distribution | Source(s) | Notes |
|---|---|---|---|---|---|
| 8 | Mortality rate among those who are not cured but on treatment, XDR-TB only, monthly | 0.01307 | Beta (mean 0.01307, s.d. 0.00436)* | Bastos M. L., et al. 2017 [4] | See Supplementary Material E "Calculated Model Parameters". |
| 9 | Probability of observed success for a fully effective WHO longer regimen, MDR-TB only (excluding XDR), at 21 months | 0.640 | Published point estimate and 95% CI (0.63-0.65) modeled as Beta (mean 0.64, s.d. 0.0051 | Bastos M. L., et al. 2017 [4] | Estimate from a large meta-analysis. The regimen duration was variable across studies, typically in the range 18-24 months. To obtain a monthly cure rate for Standard of Care strategies, we assumed a 21 month regimen duration. The published estimate is for individualized regimens; we used this parameter specifically to inform the effectiveness of a *fully effective* regimen of 4 drugs (i.e., a regimen composed of 4 drugs to which the individual's strain of *M.tb.* is truly susceptible). See also Supplementary Material E: "Calculated Model Parameters". |
| 10 | Probability of observed success for a fully effective longer regimen, XDR-TB only, 21 months | 0.26 | Published point estimate and 95% CI (0.23, 0.30) modeled as Beta (mean 0.26, s.d. 0.019 | Bastos M. L., et al. 2017 [4] | As for parameter [9]. |
| 11 | Hazard Rate Ratio of cure for each effective drug in the regimen (relative to one fewer effective drugs) | 1.65 | Published point estimate and 95% CI (1.48, 1.84) modeled as Lognormal (mu 0.5008, sigma 0.056) | Yuen, CM. et al. PLoS Med 2015 [5] | Applied to a maximum of 4 drugs (i.e., there is no further increase in the monthly cure rate for 5 drugs compared to 4). Let this parameter be *x*. Let *n* be the number of effective drugs. The individual's specific monthly cure rate under the Standard of Care is calculated by dividing the monthly cure rate for a fully effective regimen by $x^{\max(0,(4-n))}$. The referenced estimate is based on the outcome of time to sputum culture conversion. We assume the same relationship holds for the rate of true cure. |

| # | Parameter name | Point Estimate | Distribution | Source(s) | Notes |
|---|---|---|---|---|---|
| 12 | Hazard Rate Ratio of cure for the BPaLM regimen as compared to the SOC | 1.59 | Published point estimate and 95% CI (1.18, 2.14) modeled as Lognormal (mu 0.453, sigma 0.147) | Nyang'wa, B.-T. et al.. NEJM 2022 [6] | The referenced estimate is based on the outcome of time to sputum culture conversion. We assume the same relationship holds for the rate of true cure, and explore this assumption in sensitivity analysis. Longer term health outcomes such as death and failure did not provide sufficient numbers to characterize the difference in effectiveness. |
| 13 | 1 month probability of loss to follow up | Varies. See Table S2. | See "Notes" column | Walker IF et al. Eur Respir J 2019 Fig 1 [7] | See Table S2 for point estimates of the probability of LTFU each month. A measure of dispersion was calculated for each monthly probability by adopting a standard deviation equal to one third of the mean. To account for the likely dependence of the probability of LTFU between months, individuals had a percentile drawn from the standard uniform distribution (0, 1), and faced the probability each month equal to the CDF of that percentile. |
| 14 | 1 month probability of relapse (i.e. being detected and returning to treatment) for those who have active TB but were observed as successes | Month 0-29 post treatment stopping: 0<br>Month 30-98: 0.0303<br>Month 99 on: 0 | None | Blondal et al. Int J Tub Lung Dis 2012 [8] | Based on Fig 4 in the referenced paper, we assumed no relapse in the first 29 months after finishing treatment. We then assumed that there is a constant rate of return over months 30-98, and that the probability of return then drops to zero. |

| # | Parameter name | Point Estimate | Distribution | Source(s) | Notes |
|---|---|---|---|---|---|
| 15 | 1 month probability of being detected and returning to treatment for those who have been LTFU | 0.0303 | None | Blondal et al. Int J Tub Lung Dis 2012 [8] | We assumed the same rate as for those observed as success in months 30-98 after leaving treatment, but that this rate of return applies from the first month. |
| 16 | Probability of being correctly identified as a failure, among those not truly cured, at the end of each treatment cycle | 0.90 | None | Derived from other estimates | See Supplementary Material E "Calculated Model Parameters". |
| **ACQUIRED RESISTANCE** | | | | | |
| 17 | Probability of acquiring resistance to a drug, conditional on treatment with a regimen of 4 or more effective drugs, over 6 months | 0.008 | Published point estimate and 95% CI (0.005, 0.01) modeled as Beta (mean 0.008, s.d. 0.0015) | Lew W. et al. Annals Intern Med 2008 [9] | We define an "effective" drug as one to which the strain of *M. tuberculosis* is susceptible. We use the published estimate to produce a monthly rate of resistance acquisition, which is constant conditional on the number of effective drugs. |

| # | Parameter name | Point Estimate | Distribution | Source(s) | Notes |
|---|---|---|---|---|---|
| 18 | Probability of acquiring resistance to a drug, conditional on treatment with a regimen of 3 effective drugs, over 6 months | 0.060 | Published point estimate and 95% CI (0.04, 0.08) approximated and modeled as Beta (mean 0.06, s.d. 0.0102) | Lew W. et al. Annals Intern Med 2008 [9] | We define an "effective" drug as one to which the strain of *M. tuberculosis* is susceptible. We use the published estimate to produce a monthly rate of resistance acquisition, which is constant conditional on the number of effective drugs. The published estimate was for strains with "single drug resistance"; we assume this equates to a regimen with three effective drugs. |
| 19 | Increase in the rate of resistance acquisition for 2 effective drugs, compared to 3, per month | 0.009 | Lognormal (mu -4.7632, sigma 0.3246)* | Lew W. et al. Annals Intern Med 2008 [9] | Parameters [17] and [18] were converted to monthly values. We then took the difference to produce this estimate, and assume the same difference exists between 2 and 3 effective drugs, as between 3 and 4 effective drugs. |
| 20 | Probability of acquiring resistance to a drug, conditional on treatment with a regimen of 3 effective drugs, over 6 months | 0.109 | Derived from above | Derived from above | Calculated using parameters [17], [18] and [19]. Converted to a 6 month probability to facilitate comparison to parameters [17] and [18]. In Lew et al.,[9] the authors do report an estimate for a parameter *similar* to this one: the probability of acquired resistance to a new drug for those starting with a strain that is resistant to *at least* 2 of the drugs in the regimen. As we explicitly model the exact number of effective drugs at any given time, we make this a separate parameter for resistance to *exactly* two drugs. As an approximate validity check, this estimate does fall within the 95% CI for this estimate in Lew et al. (point estimate 0.14 (95% CI: [0.09, 0.2]). |

| # | Parameter name | Point Estimate | Distribution | Source(s) | Notes |
|---|---|---|---|---|---|
| 21 | 1 month probability of acquiring resistance to a new drug, conditional on starting with ONE effective drug | 0.150 | Beta(11,62) | Bulletin WHO, 1960, Table 11[10] | Estimate for standard dose isoniazid monotherapy. The authors of the referenced study reported the number of culture positive specimens each month after treatment, and the proportion of those resistant to isoniazid. We applied the proportion of resistant isolates for month 1 (i.e., 11 of 73) for each month an individual receives only one effective drug. |
| **SEVERE ADVERSE EVENTS** | | | | | |
| 22 | Probability of SAE due to pyrazinamide, 3 months | 0.028 | Beta (56,1967) | Bastos M. L., et al. 2017 Table 5 [4] | We assumed this applies to the first three months on treatment only. Converted to a one month probability in the model. |
| 23 | Probability of SAE due to ethambutol, 3 months | 0.005 | Beta(6, 1319) | Bastos M. L., et al. 2017 Table 5 [4] | As for parameter [22]. |
| 24 | Probability of SAE due to amikacin, 3 months | 0.073 | Beta(184,2354) | Bastos M. L., et al. 2017 Table 5 [4] | As for parameter [22]. Estimate is for injectables as a whole, here it is applied for amikacin. |
| 25 | Probability of SAE due to moxifloxacin, 3 months | 0.012 | Beta(10,817) | Bastos M. L., et al. 2017 Table 5 [4] | As for parameter [22]. |

| # | Parameter name | Point Estimate | Distribution | Source(s) | Notes |
|---|---|---|---|---|---|
| 26 | Probability of SAE due to thiamide, 3 months | 0.082 | Beta(173,1933) | Bastos M. L., et al. 2017 Table 5 [4] | As for parameter [22]. This estimate is applied in the model for ethionamide. |
| 27 | Probability of SAE due to cycloserine, 3 months | 0.045 | Beta(96,2044) | Bastos M. L., et al. 2017 Table 5 [4] | As for parameter [22]. |
| 28 | Probability of SAE due to linezolid, 3 months | 0.179 | Beta(140, 643) | Lan Z., et al. Lancet Resp Med 2020 [11] | We used the fixed effects estimates from the referenced paper, corresponding to a Beta distribution using the total pooled numbers of SAE and non-SAE. Applied to first three months on treatment only (by assumption). Converted to a one month probability in the model. |
| 29 | Probability of SAE due to bedaquiline, 3 months | 0.019 | Beta(9, 455) | Lan Z., et al. Lancet Resp Med 2020 [11] | As for parameter [28]. |
| 30 | Probability of SAE due to clofazimine, 3 months | 0.007 | Beta(12, 1700) | Lan Z., et al. Lancet Resp Med 2020 [11] | As for parameter [28]. |
| 31 | Probability of SAE due to isoniazid, 3 months | 0.005 | Beta(1,199) | Assumption | As for parameter [22]. |

| # | Parameter name | Point Estimate | Distribution | Source(s) | Notes |
|---|---|---|---|---|---|
| 32 | Probability of SAE due to pretomanid, 3 months | 0.025 | Beta(3, 119) | Gils et al., 2022 [12] | This systematic review presents some findings for adverse events from pretomanid monotherapy in early studies. These did not report Grade 4-5 Severe Adverse Events, but rather "Serious" Adverse Events, and we use those numbers here. We pooled the simple number of serious adverse events (n=3), and divided this by the numbers of participants across those trial arms (n=122) to provide the estimate. |
| 33 | Probability of SAE due to delamanid, 3 months | 0.008 | Beta(1,120) | Borisov S., et al. Eur Resp J 2019 [13] | As for parameter [22]. |
| **HEALTHCARE RESOURCE UTILIZATION** | | | | | |
| 34 | Number of inpatient days per typical SAE | 2 | N/A | Assumption | Assumption in line with follow-up documented by Schnippel, et al.[14] We assumed this inpatient stay takes place at a secondary care hospital. |
| 35 | Number of outpatient visits per typical SAE | 2 | N/A | Assumption | Assumption in line with follow-up documented by Schnippel, et al.[14] We assumed these visits take place at a secondary care hospital. |
| **UTILITY WEIGHTS** | | | | | |

| # | Parameter name | Point Estimate | Distribution | Source(s) | Notes |
|---|---|---|---|---|---|
| 36 | Utility weight at treatment baseline (also applied to relapse and those LTFU) | 0.750 | Published point estimate and 95% CI (0.66, 0.83) modeled as Beta (mean 0.750, s.d. 0.046) | Bauer M., et al. Qual Life Res 2015 [15] | Estimate from standard gamble, from participants from Canada. We use the estimate which was controlled for various differences between TB and non-TB groups. While we did not compare TB to no TB, this covariate adjustment may account for changes in the population over time, which helped ensure that the utility weights applied in our model were internally consistent. |
| 37 | Utility weight at 1 month of treatment | 0.900 | Published point estimate and 95% CI (0.81, 0.99) modeled as Beta (mean 0.900, s.d. 0.046) | Bauer M., et al. Qual Life Res 2015 [15] | As for parameter [36]. |
| 38 | Utility weight at 2 months of treatment | 0.890 | Published point estimate and 95% CI (0.79, 0.98) modeled as Beta (mean 0.890, s.d. 0.051) | Bauer M., et al. Qual Life Res 2015 [15] | As for parameter [36]. |
| 39 | Utility weight at 6 months of treatment | 0.920 | Published point estimate and 95% CI (0.83, 1.00) modeled as Beta (mean 0.920, s.d. 0.046) | Bauer M., et al. Qual Life Res 2015 [15] | As for parameter [36]. |
| 40 | Utility weight at 9 months of treatment | 0.970 | Published point estimate and 95% CI (0.89, 1.00) modeled as Beta (mean 0.970, s.d. 0.041) | Bauer M., et al. Qual Life Res 2015 [15] | As for parameter [36]. |

| # | Parameter name | Point Estimate | Distribution | Source(s) | Notes |
|---|---|---|---|---|---|
| 41 | Utility weight at 12 months of treatment (also applied to all those who have apparent success as soon as treatment regimen finished, if earlier than 12 months) | 1.000 | Published point estimate and 95% CI (0.92, 1.00) modeled as Beta (mean 0.99, s.d. 0.02) | Bauer M., et al. Qual Life Res 2015 [15] | As for parameter [36]. |
| 42 | Decrease in utility for severe adverse event | 0.056 | Published point estimate and S.E.M (0.006) modeled as Beta (alpha 82.18, beta 1385.27) | Takahara M, et al. Acta Diabetologica 2019 [16] | This decrement was applied as a subtraction to the base utility weight (parameters [36]-[41]) for each grade 4-5 SAE experienced, and was assumed to be lifelong. Though parameters [36]-[41] likely incorporate some disutility from treatment toxicity, we sought to capture the differences in quality of life between different drug regimens, and so we modeled an explicit decrement in utility for each grade 4-5 SAE experienced. The estimate from the referenced study was for symptomatic peripheral neuropathy among diabetic patients from participants in Japan. For parsimony, we assumed this mild but lifelong decrement reflected the average consequence among the many different possible grade 4-5 SAEs that could be experienced. |
| **COSTS (2022 USD unless otherwise specified)** | | | | | |
| 43 | Cost of treating MDR-TB, first phase (first 2 months), per month | 390.12 | Gamma (mean 390.12, s.d. 130.04)* | Chikovani I., et al. Int J Tuberc Lung Dis 2021 Table 7 [17] | From the referenced study we adopted the top-down estimates for the public sector. Referenced estimate excluded drugs and DST. Includes inpatient services as routine for the first 2 months of treatment in Georgia. Updated from 2018 GEL to 2022 USD using CPI and exchange rates from the World Bank. |
| 44 | Cost of treating MDR-TB, second phase (month 3 onwards), per month | 119.35 | Gamma (mean 119.35, s.d. 39.78)* | Chikovani I., et al. Int J Tuberc Lung Dis 2021. Table 7 [17] | Top-down estimates for public sector. Excludes drugs, DST and inpatient services (listed separately). Updated from 2018 GEL to 2022 USD using CPI and exchange rates from the World Bank. |

| # | Parameter name | Point Estimate | Distribution | Source(s) | Notes |
|---|---|---|---|---|---|
| 45 | Cost per *M. tb.* culture | 8.04 | Gamma (mean 8.04, s.d. 2.68)* | Cates L., et al. Int J Tub Lung Dis 2020. Table 1 [18] | Estimate for solid LJ medium, prepared in-house. Updated from 2018 USD to 2022 USD using exchange rates, and the CPI for Moldova from the World Bank. |
| 46 | Cost per MGIT | 20.37 | Gamma (mean 20.37, s.d. 6.79)* | Cates L., et al. Int J Tub Lung Dis 2020. Table 1 [18] | Updated from 2018 USD to 2022 USD using exchange rates, and the CPI for Moldova from the World Bank. |
| 47 | Cost per Xpert MTB/RIF | 32.57 | Gamma (mean 32.57, s.d. 10.86)* | Cates L., et al. Int J Tub Lung Dis 2020. Table 1 [18] | Updated from 2018 USD to 2022 USD using exchange rates, and the CPI for Moldova from the World Bank. |
| 48 | Cost per second-line phenotypic DST | 51.29 | Gamma (mean 51.29, s.d. 17.10)* | Cates L., et al. Int J Tub Lung Dis 2020. Table 1 [18] | Updated from 2018 USD to 2022 USD using exchange rates, and the CPI for Moldova from the World Bank. |
| 49 | Cost of amikacin treatment, per month | 35.72 | Gamma (mean 35.72, s.d. 11.91)* | Stop TB Partnership Global Drug Facility Medicines Catalog [19] | 1000mg once daily (intensive phase daily dose of 15mg/kg for a 65kg patient) |
| 50 | Cost of bedaquiline treatment, first month only | 126.70 | Gamma (mean 126.7, s.d. 42.23)* | Stop TB Partnership Global Drug Facility Medicines Catalog [19] | 400mg once daily for first 2 weeks, then 200mg 3x/week |
| 51 | Cost of bedaquiline treatment, per subsequent month | 47.12 | Gamma (mean 47.12, s.d. 15.71)* | Stop TB Partnership Global Drug Facility Medicines Catalog [19] | 200mg 3x/week |
| 52 | Cost of clofazimine treatment, per month | 19.89 | Gamma (mean 19.89, s.d. 6.63)* | Stop TB Partnership Global Drug Facility Medicines Catalog [19] | 100mg once daily |

| # | Parameter name | Point Estimate | Distribution | Source(s) | Notes |
|---|---|---|---|---|---|
| 53 | Cost of cycloserine treatment, first month only | 21.95 | Gamma (mean 21.95, s.d. 7.32)* | Stop TB Partnership Global Drug Facility Medicines Catalog [19] | 250mg twice daily for first 2 weeks, then 500mg twice daily |
| 54 | Cost of cycloserine treatment, per subsequent month | 28.52 | Gamma (mean 28.52, s.d. 9.51)* | Stop TB Partnership Global Drug Facility Medicines Catalog [19] | 500mg twice daily |
| 55 | Cost of delamanid treatment, per month | 307.62 | Gamma (mean 307.62, s.d. 102.54)* | Stop TB Partnership Global Drug Facility Medicines Catalog [19] | 100mg twice daily |
| 56 | Cost of ethambutol treatment, per month | 3.35 | Gamma (mean 3.35, s.d. 1.12)* | Stop TB Partnership Global Drug Facility Medicines Catalog [19] | 1200mg once daily (recommended dose for 56-75 kg patient).[20] |
| 57 | Cost of ethionamide treatment, per month | 11.16 | Gamma (mean 11.16, s.d. 3.72)* | Stop TB Partnership Global Drug Facility Medicines Catalog [19] | 500mg twice daily (approximately 15mg/kg for 65kg patient) |
| 58 | Cost of isoniazid treatment, per month | 0.57 | Gamma (mean 0.57, s.d. 0.19)* | Stop TB Partnership Global Drug Facility Medicines Catalog [19] | 300mg once daily |
| 59 | Cost of linezolid treatment, per month | 22.44 | Gamma (mean 22.44, s.d. 7.48)* | Stop TB Partnership Global Drug Facility Medicines Catalog [19] | 600mg twice daily |
| 60 | Cost of moxifloxacin treatment, per month | 4.86 | Gamma (mean 4.86, s.d. 1.62)* | Stop TB Partnership Global Drug Facility Medicines Catalog [19] | 400mg once daily |

| # | Parameter name | Point Estimate | Distribution | Source(s) | Notes |
|---|---|---|---|---|---|
| 61 | Cost of pretomanid treatment, per month | 60.80 | Gamma (mean 60.8, s.d. 20.27)* | Stop TB Partnership Global Drug Facility Medicines Catalog [19] | 200mg once daily. |
| 62 | Cost of pyrazinamide treatment, per month | 1.86 | Gamma (mean 1.86, s.d. 0.62)* | Stop TB Partnership Global Drug Facility Medicines Catalog [19] | 1500mg once daily (recommended dose for 56-75kg patient).[21] |
| 63 | Cost per inpatient bed day | 92.71 (2018 GEL) | Gamma (mean = 92.71, s.d. = 7.83) | Value TB Dataset [22] | MDR-specific estimate for an urban tertiary hospital. Converted to 2022 USD in model. |
| 64 | Cost per outpatient visit | 6.49 (2018 GEL) | Gamma (mean = 6.49, s.d. = 1.21) | Value TB Dataset [22] | Estimate for adherence support visit for an urban tertiary hospital. Converted to 2022 USD in model. |
| 65 | Direct nonmedical costs of receiving RR-TB treatment, 18 months | 2044.46 | Simulated from bootstrapped empirical estimates (mean 2044.46, 95% UI (1354.06, 2999.25)) | Portnoy A. et al., Lancet Glob Health 2023, Table S9 [23] | Applied to all individuals undergoing TB treatment. Values were drawn from this distribution and then converted to a monthly cost, assuming 18 months of TB treatment in Moldova. |
| 66 | Indirect cost of having RR-TB, 18 months | 2301.13 | Simulated from bootstrapped empirical estimates with (mean 2301.13, 95% UI (1359.23, 3744.06)) | Portnoy A. et al., Lancet Glob Health 2023, Table S10 [23] | Applied to all individuals with TB disease, whether they were receiving treatment or not, and also to those truly cured but yet to complete the treatment course. Values were drawn from this distribution and then converted to a monthly cost, assuming 18 months of TB treatment in Moldova. |
| 67 | GDP per capita for Moldova, per year (2022 USD) | 5529 | N/A | World Bank national accounts data [24] | This is the GDP per capita for Moldova estimated using current prices. |
| 68 | Final consumption expenditure, as a proportion of GDP (2021) | 0.987 | N/A | World Bank national accounts data [24] | Assumed same value for 2022 as for 2021. The complement of this number (0.013) is multiplied by the monthly GDP of Moldova to provide the cost per month of premature death (i.e. postfatal productivity loss). |

| # | Parameter name | Point Estimate | Distribution | Source(s) | Notes |
|---|---|---|---|---|---|
| 69 | Cost of conducting LTFU tracing, per episode | 4.96 (2018 GEL) | Gamma (mean 4.96, s.d. 1.91) | Value TB Dataset [22] | Estimate for community-based LTFU tracing over the phone. Converted to 2022 USD in model. |
| 70 | Life Expectancy, Moldova | 71.2 years | N/A | UN Data World Population Prospects [25] | Estimate for individuals aged 42, the average age of the cohort, in 2021. Used only to calculate productivity losses for all those who die prior to this age. |
| 71 | GDP Deflator Indices | Moldova: 100 (2010), 108.2 (2011), 120.8 (2013), 140.7 (2015), 158.0 (2017), 181.2 (2020), 187.2 (2021), 193.2 (2022)*; Georgia: 116.19 (2018), 131.12 (2020), 144.6 (2021), 158.1 (2022)* | N/A | World Bank GDP Deflator [24] | Applied to account for inflation. *2022 values are not yet available, and were calculated under the assumption that GDP deflator increased by the same amount from 2021 to 2022 as from 2020 to 2021. |
| 72 | US Dollar to Moldovan Leu (USD/MDL) market exchange rates | 13.84 (2014), 16.79 (2018), 17.33 (2020), 18.78 (2022) | N/A | Xe.com [26] | Annual mean calculated by averaging monthly values. |
| 73 | US Dollar to Georgian Lari (USD/GEL) market exchange rates | 2.53 (2018), 3.09 (2020), 2.95 (2022) | N/A | Xe.com [26] | Annual mean calculated by averaging monthly values. |

| # | Parameter name | Point Estimate | Distribution | Source(s) | Notes |
|---|---|---|---|---|---|
| 74 | US Dollar to Moldovan Leu (USD/MDL) PPP exchange rate | 2.558 (2011), 3.217 (2017) | N/A | World Bank International Comparisons Project [27] | Estimates for "Actual Health". |
| 75 | US Dollar to Georgian Lari (USD/GEL) PPP exchange rate | 0.413 (2011), 0.461 (2017) | N/A | World Bank International Comparisons Project [27] | Estimates for "Actual Health". |
| 76 | Willingness-to-pay for improvements in health (2015 USD) | Lower estimate: 85% GDP per capita Upper estimate: 127% GDP per capita | N/A | Ochalek et al., 2018 [28] | Applied to 2022 GDP per capita, providing a lower estimate of 4699; upper estimate 7021 in 2022 USD. |
| 77 | Number of individuals with RR-TB initiating treatment in Moldova, per year | 644 | Normal (mean 644, s.d. 215)* | WHO Global TB Programme, Case notifications [29] | Used to estimate budget impact only. Calculated as the annual mean of RR-TB (i.e., inclusive of MDR, pre-XDR, XDR) case notifications reported to the WHO over the five year period 2018-2022, inclusive. Data reporting changed over this period. For years 2018 and 2019, we summed the reported numbers for RR/MDR-TB (which were exclusive of XDR) and XDR-TB. For 2020-2022 we added the number of laboratory confirmed RR-TB with fluoroquinolone resistance to the number of RR-TB without known fluoroquinolone resistance. |
| **TEST CHARACTERISTICS** | | | | | |
| 78 | MGIT DST Sensitivity for amikacin resistance | 87.5% | Beta(7,1) | Tekin K., et al. 2017 [30] | In the referenced study, 6 resistant samples were identified using the gold standard. All were correctly identified by MGIT. These values were used to update a Beta(1,1) prior. |

| # | Parameter name | Point Estimate | Distribution | Source(s) | Notes |
|---|---|---|---|---|---|
| 79 | MGIT DST Specificity for amikacin resistance | 97.62% | Beta(41,1) | Tekin K., et al. 2017 [30] | In the referenced study, 40 susceptible samples were identified using the gold standard. All were correctly identified by MGIT. We used these values used to update a Beta(1,1) prior. |
| 80 | MGIT DST Sensitivity for fluoroquinolone resistance | 77.78% | Beta(20,1) | Devasia R. A., et al. 2009 [31] | In the referenced study, 19 resistant samples were identified using the gold standard. All were correctly identified by MGIT. We used these values used to update a Beta(1,1) prior. |
| 81 | MGIT DST Specificity for fluoroquinolones resistance | 97.56% | Beta(779,1) | Devasia R. A., et al. 2009 [31] | In the referenced study, 778 susceptible samples were identified using the gold standard. All were correctly identified by MGIT. We used these values used to update a Beta(1,1) prior. |

CDF – cumulative distribution function; CI – confidence interval; CrI – credibility interval; CPI – Consumer Price Index; DST – drug susceptibility testing; LJ – Lowenstein-Jensen; MDL – Moldovan Leu; GDP – Gross Domestic Product; GEL – Georgian Lei; LTFU – lost to follow up; M. tb. – mycobacterium tuberculosis; NHB – Net Health Benefit; QALY – Quality-adjusted Life Year; SAE – Severe Adverse Event; SEM – Standard Error of the Mean; SMR – standardized mortality ratio; UI – Uncertainty Interval; USD – United States Dollars; WTP – Willingness-to-Pay.

*denotes a parameter where there was no readily available measure of dispersion. For these parameters, we assumed a standard deviation equal to one third of the mean.

**REFERENCES**

These references are provided here for convenience. They are also provided within the main manuscript file in the legend for S1 Table.

1. World Population Prospects. UN Population Division, Department of Economic and Social Affairs.; 2019. Available: https://population.un.org/wpp/Download/Files/1_Indicators%20(Standard)/EXCEL_FILES/3_Mortality/WPP2019_MORT_F15_1_LIFE_TABLE_SURVIVORS_BOTH_SEXES.xlsx

2. Ragonnet R, Flegg JA, Brilleman SL, Tiemersma EW, Melsew YA, McBryde ES, et al. Revisiting the Natural History of Pulmonary Tuberculosis: A Bayesian Estimation of Natural Recovery and Mortality Rates. : 9.

24. World Bank Open Data. Available: https://data.worldbank.org/

25. UNdata | record view | Life expectancy at birth for both sexes combined (years). [cited 24 Mar 2022]. Available: http://data.un.org/Data.aspx?q=moldova&d=PopDiv&f=variableID%3A68%3BcrID%3A498

26. xe.com Currency Charts. Available: https://www.xe.com/currencycharts/
