## Supplemental Appendix 1 for "Impact and cost-effectiveness of the 6-month BPaLM regimen for rifampicin-resistant tuberculosis: a mathematical modeling analysis"

**S1 Appendix. Additional Detail on Microsimulation Model**

1. *Markov Health States*

The model mechanisms included Markov health states and individual patient- and drug-level trackers. The Markov health states were:

(1): Receiving TB treatment

(2): TB disease not receiving treatment

(3): Cured post-treatment

(4): Dead

2. *Transitions between Markov Health States*

The Markov cycle length was one month. All individuals started in state (1), and stayed there until the earliest of: death, LTFU, or discharge. While in state (1), an individual could be cured of TB disease. We assumed that all truly cured patients were correctly identified as such; these individuals were discharged after the completion of their planned regimen and transitioned to state (3). When those individuals not truly cured were assessed at the end of treatment, 90% (Table S1) received a 6 month treatment extension, and the remainder were incorrectly assessed to have successfully completed treatment and were discharged. Patients receiving treatment extensions were assessed in the same way at the end of each extension. Individuals discharged before true cure were moved into state (2) and faced a monthly probability of relapsing and returning to treatment.

Individuals also faced a risk each month of becoming LTFU; if this occurred before they were cured, they transitioned to state (2) with a monthly probability of returning to treatment, and if they had already been truly cured they moved into state (3). Self-cure was possible from state (2), at a lower rate than cure on treatment. All simulated individuals faced a risk of death each month, with the mortality rate lower following cure. Prior to cure, the rate of death was higher for those who had stopped treatment. Quality of life was lowest at the start of treatment, and improved as the treatment course progressed, until it reached the maximum value upon true cure.

#### *3. Individual patient- and drug-level trackers*

For each simulated individual we tracked whether they had been truly cured during treatment (i.e., before they reached the end of the prescribed regimen and moved into state (3)), their current drug regimen, the duration of treatment with each drug, whether the strain of *M. tuberculosis* was resistant to each drug, the duration of resistance to each drug since treatment initiation, the results of the most recent DST performed, whether they had experienced a SAE to each drug, and the observed EOT outcomes (Fig S6). This set of trackers collectively informed the event probabilities and health state utility weights (Table S1). In each month, these trackers were also used to calculate the number of effective drugs, by matching the drugs being used in the treatment regimen with the (true) resistance profile to that drug. This variable influenced the probability of cure and the probability that each individual's strain of *M. tuberculosis* would acquire resistance to any new drugs (Table S1, Fig S4). Differential adherence by strategy was not modeled explicitly, and we assumed that the effects of imperfect adherence were reflected in published effectiveness estimates.

#### *4. Outcomes*

##### *4.1 Calculating Net Health Benefit*

NHB was calculated according to convention [1]: total discounted costs were converted into QALYs of equivalent value using the exchange rate of WTP for gains in health, itself measured in $ per QALY. These were then subtracted from the total discounted QALYs to produce NHB, measured in units of QALYs.

*4.2 End of Treatment Outcomes*

We recorded the observed EOT outcome for each simulated individual in the model. We then validated these modeled outcomes against data reported to the WHO from Moldova, presented separately for RR-TB (i.e., MDR/RR-TB) overall and for the subset of patients with XDR-TB. In this analysis, the definition of XDR-TB used is the older definition used by WHO [2], meaning TB that is resistant to any fluroquinolone and to at least one of three second-line injectable drugs, in addition to isoniazid and rifampicin. We used the older definition to allow validation of the modelled EOT outcomes against WHO data. The classification structure for recording EOT outcomes in the model is shown in Fig S6, along with our best interpretation of the EOT structure in the empirical data.

*4.3 Calculating the Budget Impact*

We tracked the subset of direct medical outcomes borne by Moldova's national TB program, and tallied these for each individual in each cycle under the following categories: Drugs, laboratory tests, routine inpatient and outpatient care, and non-routine inpatient and outpatient care (i.e., stemming from the treatment of grade 4-5 SAEs, for adjustment of a regimen following the detection of resistance on DST, or for LTFU tracing). We ran the model for 12, 24, 36, 48, and 60 months with a constant randomization seed. We used these results to calculate the average budget impact for years 1 through 5 for an individual starting treatment at the beginning of year 1, then repeated this process for cohorts starting treatment at the beginning of years 2, 3, 4, and 5 to account for all the patients who may be treated over a five year period.

We built a separate model in R to scale up from the budget impact of an individual to that of a policy change for the Moldovan national TB program. To obtain the number of case notifications per year, we averaged over WHO RR-TB case notifications for the period 2018-2022 [3] (Table S1). We then divided this number by 12 to estimate the number of case notifications per month. We accounted for the budget impact for each monthly starting cohort, from the time they started treatment to the end of the five year budget impact horizon, and summed across all monthly starting cohorts to estimate the budget impact to the national TB program.

### *5. Modeled events*

#### *5.1 Death*

Patients were exposed to a monthly risk of mortality, incorporating the risk from background causes and disease-specific mortality from TB. The TB-specific mortality risk was highest for individuals with TB disease that was not currently being treated. Treatment lowered the TB-specific mortality risk. After an individual was cured, the TB-specific mortality risk decreased but was still greater than zero to account for post-TB sequelae (Table S1).

#### 5.2 *Cure*

For the first month of treatment and for untreated TB disease, an individual was allowed to self-cure. A higher monthly cure rate was assumed from month two of treatment onwards (conditional on the treatment strategy and the number of effective drugs each month, as previously mentioned).

#### *5.3 Loss to follow-up (LTFU)*

Each month, an individual receiving treatment could become LTFU, with a decreasing monthly probability over time (Table S2). Those LTFU before true cure entered the “2) TB Disease – no longer receiving treatment” state and were allowed to subsequently recommence treatment. Those LTFU *after* true cure, but while still receiving treatment, entered the “3) Cured post-treatment” state and did not return to treatment. The probability of return to treatment is described in Table S1.

#### *5.4 Drug resistance acquisition*

The resistance status of each individual's strain of *M. tuberculosis* to each drug was assumed to be binary (susceptible or resistant). The probability of developing resistance in a given month to each drug was a function of the number of effective drugs in the regimen. Conditional on the number of effective drugs, the probability of acquiring resistance was independent and identically distributed for all drugs to which *M. tuberculosis* was exposed in any month, with three exceptions: pretomanid resistance was assumed to confer immediate delamanid resistance and vice versa [4], bedaquiline resistance was assumed to confer immediate clofazimine resistance (but not vice versa) [5], and the rate of acquiring resistance to linezolid was assumed to be half the rate as to other drugs [6,7].

We assumed that if an individual had resistance to a drug at a given time, the *M. tuberculosis* strain could not later revert to being susceptible. While resistance status was tracked for all patients with TB disease regardless of whether they were being treated, the probability of acquiring resistance fell to zero for all drugs in the month following treatment cessation.

#### *5.5 Drug Susceptibility Testing*

Individuals received DST by MGIT for moxifloxacin and amikacin according to the frequency prescribed by the strategy (Table 1). We assumed that individuals who had been truly cured would not be able to produce an adequate sputum sample, and as such did not undergo DST. We also assumed full adherence to the prescribed DST regimen. In the event of detecting new drug resistance, the respective drug was discontinued for that individual with a lifetime contraindication, and was replaced immediately. The only exception to this rule of replacement was for moxifloxacin under variations of the 6 months of BPaLM strategy, where some strategies continued BPaL only, rather than replacing moxifloxacin (Table 1). Sensitivity and specificity values for DST were estimated from the literature (Table S1).

#### *5.6 Severe Adverse Events (SAEs)*

We modeled a generalized severe (i.e., grade 4-5) treatment-related SAE, which we assumed took place in any of the first three months of treatment with each drug, but not after. An SAE resulted in the responsible drug being discontinued that month, with a lifelong contraindication and small lifelong decrement to that individual's health-related quality of life (Table S1). Each additional SAE was assumed to confer the same incremental reduction in quality of life and healthcare resource utilization.

#### 6. *Missing Data*

In the genomic sequencing dataset, for observations with missing age (n = 12; 1.8%) we imputed the mean age of the cohort (42 years). There was no missing drug resistance data.

#### 7. *Probabilistic Sensitivity Analysis (PSA)*

For each input parameter, distributions were fit to the published measure of dispersion (95% confidence intervals or standard deviation) where available. Where there was no accompanying measure of dispersion, we assumed a standard deviation equal to one third of the mean. Information on all distributions is provided in Table S1. PSA results were obtained by performing $2^{nd}$-order Monte Carlo simulation with 1,000 iterations. In each iteration, a set of values was drawn from the parameter distributions and the "inner loop" (i.e., 10,000 $1^{st}$-order Monte Carlo simulations) was run. 95% uncertainty intervals (UIs) were calculated as the $2.5^{th}$ and $97.5^{th}$ centiles from the PSA results. Due to the challenges in interpreting negative ICERs [1,8], we did not provide 95% UIs for ICERs.

The p-values in this study were constructed from the PSA results. As in other health economic evaluation using simulation models [9], we provide two-tailed p-values as two times the proportion of PSA iterations that would lead to the opposite direction of effect. We did this for all difference metrics (e.g., incremental QALYs and incremental costs, difference in proportion with an unfavorable outcome) by constructing an empirical cumulative distribution function (CDF) from the PSA results. For distributions where the median difference is greater than 0, we provided the p-value as the two times the CDF of 0 (i.e.,

2*CDF(0)), and for distributions where the median difference is less than 0, we provided the p-value as two times the complement (i.e., 2*(1-CDF(0))).
